# Exploratory Blood Biomarker Patterns in a Mixed Dementia Cohort

**DOI:** 10.1101/2024.11.07.24316927

**Authors:** Haşim Gezegen, Merve Alaylıoğlu, Erdi Şahin, Owen Swann, Elena Veleva, Gamze Güven, Umran Yaman, Derviş A. Salih, Başar Bilgiç, Haşmet Hanağası, Hakan Gürvit, Murat Emre, Duygu Gezen-Ak, Erdinç Dursun, Henrik Zetterberg, John Hardy, Amanda Heslegrave, Maryam Shoai, Bedia Samancı

## Abstract

Alzheimer’s disease (AD) diagnosis is challenging due to overlapping symptoms with other dementias. Current diagnostic methods are invasive and costly, highlighting the need for accessible biomarkers. This study investigates the diagnostic performance and pathophysiological implications of a novel plasma biomarker panel in a mixed dementia cohort, aiming to enhance diagnosis and elucidate underlying pathogenic mechanisms.

120 plasma biomarkers were analyzed using the NULISA™ platform in a well-characterized mixt dementia. CSF biomarkers were measured via ELISA. Statistical analyses employed ANOVA, and Kruskal-Wallis tests for group comparisons. Spearman correlations assessed relationships between CSF and plasma biomarkers. Diagnostic accuracy was evaluated using regression models and ROC curves. Feature importance and selection were performed using random forest analysis. Protein interactions assessed with GO enrichment analysis.

We evaluated 248 subjects (130 females, 118 males) with 117 AD, 50 MCI, 39 FTD, 25 DLB, and 17 other dementias. Plasma pTau were significantly elevated in AD compared to other groups, and in DLB compared to MCI. Plasma Aβ42 was highest in DLB, while NfL was highest in FTD. Plasma GFAP was highest in AD and elevated in DLB compared to MCI and FTD.

Plasma pTau levels showed a negative correlation with CSF Aβ42 and a positive correlation with CSF pTau in the entire cohort. CSF and plasma NfL levels were also highly correlated. These correlations were stronger in DLB and amyloid-positive MCI groups but weaker or absent in the AD group. Plasma pTau, GFAP, and NfL were negatively correlated with MMSE in AD, while GFAP showed a negative correlation with MMSE in FTD and DLB.

Plasma pTau217 demonstrated the best diagnostic accuracy for AD, DLB, FTD diagnosis and CSF amyloid positivity (AUCs 0.9, 0.84, 0.79, and 0.87, respectively). pTau181, pTau217, pTau231, total-tau and GFAP had lower odds in DLB and FTD compared to AD. AGRN, CXCL1, SCNB, TEK, and UCHL1 had higher odds in DLB compared to AD. SNAP25 had lower odds ratio in FTD compared to AD and DLB. pTau181, pTau217, pTau231, GFAP, MAPT, SNAP25 and PGF is related to AD progression.

Random forest analysis incorporating all plasma biomarkers, age, and gender yielded an AUCs of 0.85 for AD, 0.84 for FTD, and 0.75 for DLB. Refining the model by including biomarkers identified as significant in regression model improved performance, resulting in AUCs of 0.88 for AD, 0.87 for FTD, and 0.81 for DLB. This demonstrates potential for enhancing diagnostic accuracy through targeted biomarker panel refinement.

## Introduction

Alzheimer’s disease (AD) is the most prevalent age-related neurodegenerative dementia, affecting 60-80% of dementia patients and posing a significant threat to the aging population. Other common dementia types include dementia with Lewy bodies (DLB) and frontotemporal dementia (FTD), which often exhibit overlapping symptoms and pathology with AD ^1^, leading to misdiagnosis rates as high as 30% ^2^. Early and accurate diagnosis is crucial for effective disease management and therapeutic intervention. However, the current diagnostic landscape for AD and related dementias is complex. Clinical diagnosis based on symptoms alone is often inaccurate, with up to a third of clinically diagnosed AD cases lacking AD neuropathological changes post-mortem ^2^ highlighting the need for reliable biomarkers.

Cerebrospinal fluid (CSF) and neuroimaging biomarkers are valuable tools in diagnosing AD. Established AD CSF biomarkers like Aβ42 (or Aβ42/40), phosphorylated tau (pTau) and total tau (tTau) reflect key aspects of AD pathology, such as senile plaque pathology and neurofibrillary tangle formation, and axonal degeneration ^3^. Although valuable for early AD diagnosis and clinical settings ^4^, these markers inadequately capture AD’s biological diversity and struggle to differentiate AD from other dementias with overlapping pathologies ^5,6^. Comprehensive CSF proteome analysis offers the potential to reveal novel markers, providing a deeper understanding of AD’s complex pathophysiology and aiding in the identification of therapeutic targets ^7^.

Diagnostic challenges also extend to other neurodegenerative diseases like DLB. α-synuclein pathology, a hallmark of DLB, is also present in over 40% of AD cases, complicating differentiation ^8^. Core CSF biomarkers for AD show limited accuracy in distinguishing DLB from AD due to frequent co-occurrence of AD pathology in DLB patients ^9–11^. Identifying co-pathologies is crucial as they can worsen clinical outcomes. In DLB/AD co-pathology, this manifests as a more severe clinical presentation with decreased memory, heightened delusions and hallucinations, and accelerated disease progression ^12,13^. Co-pathologies with FTD are possible even in mixed AD and DLB patients, exacerbating neurodegeneration and complicating diagnosis and treatment ^14^.

AD-related pathophysiological changes can be accurately detected through various methods: CSF analysis for alterations in Aβ42, pTau, and tTau levels; PET imaging for amyloid plaque and tau neurofibrillary tangle distribution; and structural MRI for brain atrophy assessment ^15^. However, their widespread use in primary care is limited by invasiveness (CSF), radiation/cost (PET), and accessibility/cost-effectiveness, especially in resource-limited settings where dementia incidence is rising ^16–18^. This lack of reliable and accessible biomarkers poses a significant challenge, made more urgent by the emergence of disease-modifying treatments.

Plasma biomarkers offer a promising avenue for improving AD diagnosis, disease understanding, and clinical management ^16^. These biomarkers could facilitate the evaluation of therapeutic interventions and enable the assessment of future disease risk in asymptomatic individuals, as well as longitudinal monitoring of symptomatic patients ^19^. Compared to CSF or neuroimaging biomarkers, they have the potential to be simpler, more cost-effective, and easier to implement ^20,21^. Plasma pTau and amyloid biomarkers, in particular, have shown strong associations with AD pathology and cognitive decline ^22–24^. Additionally, plasma GFAP and NfL have emerged as potential biomarkers for AD and other neurodegenerative diseases, further expanding the potential of blood-based diagnostics ^25,26^.

The initiation of monoclonal antibody therapy for early AD, such as lecanemab, requires confirmation of AD pathology via biomarker testing, reflecting a shift towards prevention and preclinical treatment ^27^. Given the high AD prevalence, increasing demand for therapies, and the need for clinical trials, innovative models for connecting individuals with resources are crucial. Routine blood draws as a primary care triage tool for anti-amyloid therapy hold promise for expedited diagnosis, treatment access, and clinical trial enrolment. Despite current limitations, the pursuit of AD blood-based biomarkers remains vital for enhanced screening, diagnosis, and disease management.

NUcleic acid Linked Immuno-Sandwich Assay (NULISA), a platform using antibody-based measurements with a sequencing output, offers advantages such as low sample volume requirements and high multiplexing capacity ^28^. Its CNS panel encompasses traditional AD biomarkers, synaptic, microglial, neuroinflammation, and emerging neurodegeneration biomarkers, and has demonstrated effectiveness in biomarker detection and accuracy compared to traditional immunoassays ^29^.

In this study, we used NULISA™ to analyse 120 plasma biomarkers across diverse biological mechanisms in a well-characterized cohort, including patients with mild cognitive impairment (MCI), AD, DLB, and FTD from a single referral memory center. The study had two main objectives: First, to evaluate this novel technology, validate known diagnostic biomarkers, and identify new ones that could improve diagnostic accuracy. Second, to uncover novel plasma proteomic alterations driving the pathogenesis of AD, DLB, and FTD, thereby identifying promising new biomarker candidates for future research.

## Material and methods

### Patients

Patients were recruited from Behavioural Neurology and Movement Disorders Unit, Department of Neurology, Istanbul Faculty of Medicine, Istanbul University, a referral center for cognitive disorders. The study was conducted in accordance with the ethical standards of the Declaration of Helsinki and approved by the Istanbul University Ethics Committee (approval number 2023/1191, granted 22/06/2023).

The study encompassed a comprehensive assessment of participants, including medical history, physical and neurological examinations, psychiatric screening, clinical laboratory tests, and, cranial magnetic resonance imaging. The neuropsychological battery included the Mini-Mental Status Examination ^30^ and the Clinical Dementia Rating Scale ^31^. Also a subset of patients completed a comprehensive neuropsychological assessment at baseline.

Experienced clinicians made diagnoses based on established criteria ^15,32–35^, which included progressive cognitive complaints corroborated by an informant, clinically observed cognitive impairment as previously described, clinical laboratory tests and MRI findings that excluded other potential causes of impairment as well as CSF draw. Genetic causes were excluded for the FTD group.

Inclusion criteria for the study were age 55 years or older and signed informed consent for biological sample extraction and storage. Exclusion criteria included major systemic or psychiatric illness, significant sensory deprivation, and contraindications for the required complementary tests. Patients who underwent CSF examination between June 23, 2023, and March 25, 2024, and met the inclusion criteria were included in the study.

### CSF Biomarkers and NULISA Measurements

All participants underwent a lumbar puncture (LP) to measure Aβ42, pTau181, tTau, and NfL levels, as well as blood sample collection.

Following an overnight fast, LP was performed at 09.00 am using a 25-G needle. Ten mL of CSF, collected in four polypropylene tubes, was kept on ice for up to one hour before centrifugation at 2000 g for 10 minutes at 4°C. Previously mentioned biomarkers were run on the same day. Aliquots (250 μL) were stored in 1-mL polypropylene tubes at -80°C until analysis.

Blood samples, collected concurrently with LP, were processed within one hour. Plasma was obtained by collecting 10 mL of blood into EDTA tubes, inverting five times, and centrifuging at 2000 g for 10 minutes at 4°C. Aliquots (250 μL) were stored in 1-mL polypropylene tubes at -80°C.

The CSF levels of Aβ42, pTau181, tTau, and NfL were measured at Brain and Neurodegenerative Disorders Laboratory, Department of Neuroscience, Institute of Neurological Sciences, Istanbul University-Cerrahpasa, which is an active member of The Alzheimer’s Association Quality Control (AAQC) program ^36^. The measurements were done with INNOTEST β-AMYLOID (1-42) ELISA kit (81576, FUJIREBIO), INNOTEST PHOSPHO-TAU (181P) ELISA Kit (81574, FUJIREBIO), INNOTEST hTAU Ag ELISA Kit (81572, FUJIREBIO) and NF-light ELISA kit (10-7001, UmanDiagnostics) respectively. All measurements were performed according to the manufacturer’s assay protocol, and each sample and standard were tested in duplicate.

Plasma biomarkers were measured using the NULISA platform, developed by Alamar Biosciences, following the protocol established in ^28^.

### Statistical analysis

All analyses were performed in R 4.2.2. Data were log2-transformed and tested for normality (NPQ values from NULISA were already log_2_ transformed and no additional transformation was used). One-way ANOVA or Welch’s ANOVA was used for normally distributed data, and Kruskal-Wallis test for non-normally distributed data. Head-to-head comparisons between CSF and plasma biomarkers were performed using Spearman correlations with FDR correction. Scatter plots with linear regression lines investigated the relationship between CSF and plasma biomarkers.

To evaluate diagnostic accuracy, the dataset was divided into training and testing sets. A repeated 10-fold cross-validation was employed. All models adjusted for age and gender. Predictive performance comparison for biomarkers were performed by using ROC curves and calculating the AUC. Differences between AUC values were tested with the DeLong test. Cut-off values maximising Youden indices were used for sensitivity and specificity. The analysis was conducted with *caret* and *pROC* packages.

Multinomial logistic regression assessed the association between plasma biomarkers and diagnostic categories. A separate model was fitted for each biomarker. Odds Ratios (OR) were calculated to quantify the likelihood of a specific diagnosis relative to the biomarker levels. Adjusted p-values were computed using the FDR method. We performed an age and sex adjusted ordinal regression model to assess the associations between plasma biomarkers and disease stage in the AD continuum. These models were conducted using *nnet* and *MASS* packages.

Random forest analyses were conducted with all biomarkers and biomarkers that differed between diagnosis based on multinomial logistic regression and calculated the importance of all features. All features were tested with ten-fold cross-validation and mean AUC was used as the performance metric. The model was trained with the method argument set to “*rf*”. The analysis was conducted using *caret* package.

The Gene Ontology (GO) enrichment analyses were performed on multinomial logistic regression results, and the findings were visualized with the *clusterProfiler* package.

## Results

### Clinical and Demographical Features

We analysed data from 248 subjects (130 females, 118 males) with a mean age of 66.7 (± 9.1) years and an MMSE of 21.7 (± 7.1). Diagnoses included 117 AD, 50 MCI, 39 FTD, 25 DLB, and 17 other dementias. Other dementias consisted of NPH (normal pressure hydrocephalus), CBS (corticobasal syndrome), PSP (progressive supranuclear palsy) and MND (motor neuron disease) without dementia. **Table 1** details MMSE, CDR, and CSF amyloid (A), tau (T), and neurodegeneration (N) group distribution by diagnosis, showing no gender differences. The DLB and FTD groups differ from other groups for age and the MCI group differed from other groups for MMSE. **Table 2** presents demographics for AD continuum patients.

**Table 1.** Demographics of the cohort.

| Variable | Overall<br>N = 248 | AD<br>N = 117 | MCI<br>N = 50 | FTD<br>N = 39 | DLB<br>N = 25 | Other<br>N = 17 | p-value<sup>1</sup> |
| --- | --- | --- | --- | --- | --- | --- | --- |
| Age, mean (SD) | 66.7 (9.1) | 66.5 (8.5) | 66.2 (9.6) | 61.9 (8.6) | 74.7 (5.6) | 68.9 (8.9) | <0.001 |
| Female n (%) | 118 (48%) | 53 (45%) | 25 (50%) | 16 (41%) | 16 (64%) | 8 (47%) | 0.4 |
| MMSE, mean (SD) | 21.7 (7.1) | 20.4 (7.1) | 27.5 (2.0) | 19.8 (7.9) | 20.3 (6.5) | 19.3 (8.9) | <0.001 |
| CDR, range (median) | 0-3 (0.5) | 0-3 (0.5) | 0-0.5 (0.5) | 0-3 (1) | 0.5-2 (0.5) | 0.5-2 (1) |  |
| ATN, n (%) |  |  |  |  |  |  |  |
| A-T-N- | 46 (19%) | 0 (0%) | 20 (40%) | 12 (31%) | 9 (36%) | 5 (29%) |  |
| A-T-N+ | 5 (2.0%) | 0 (0%) | 2 (4.0%) | 3 (7.7%) | 0 (0%) | 0 (0%) |  |
| A-T+N- | 11 (4.4%) | 0 (0%) | 8 (16%) | 1 (2.6%) | 1 (4.0%) | 1 (5.9%) |  |
| A-T+N+ | 27 (11%) | 0 (0%) | 12 (24%) | 10 (26%) | 2 (8.0%) | 1 (5.9%) |  |
| A+T-N- | 40 (16%) | 5 (4.3%) | 5 (10%) | 11 (28%) | 10 (40%) | 9 (53%) |  |
| A+T-N+ | 2 (0.8%) | 1 (0.9%) | 0 (0%) | 0 (0%) | 1 (4.0%) | 0 (0%) |  |
| A+T+N- | 7 (2.8%) | 5 (4.3%) | 2 (4.0%) | 0 (0%) | 0 (0%) | 0 (0%) |  |
| A+T+N+ | 110 (44%) | 106 (91%) | 1 (2.0%) | 2 (5.1%) | 2 (8.0%) | 1 (5.9%) |  |

AD: Alzheimer dementia, MCI: Mild cognitive impairment, FTD: Frontotemporal dementia, DLB: Dementia with Lewy bodies, Other: NPH (normal pressure hydrocephalus), CBS (corticobasal syndrome), PSP (progressive supranuclear palsy) and MND (motor neuron disease) without dementia
<sup>1</sup>Kruskal-Wallis rank sum test; Pearson's Chi-squared test

**Table 2.** Demographics of AD Stages.

| Variable | Overall<br>N = 102 | Stage 2<br>N = 8 | Stage 3<br>N = 15 | Stage 4<br>N = 34 | Stage 5<br>N = 31 | Stage 6<br>N = 14 | p-value<sup>1</sup> |
| --- | --- | --- | --- | --- | --- | --- | --- |
| Age, mean (SD) | 66.1 (9.0) | 67.6 (13.1) | 69.7 (6.0) | 66.7 (8.4) | 65.4 (9.7) | 61.5 (8.0) | 0.12 |
| Gender, female (%) | 53 (52%) | 2 (25%) | 9 (60%) | 17 (50%) | 19 (61%) | 6 (43%) | 0.4 |
| MMSE, mean (SD) | 21.0 (7.1) | 28.3 (2.1) | 28.2 (1.3) | 24.6 (2.8) | 18.0 (1.9) | 8.2 (3.8) | <0.001 |
| CDR, range (median) | 0-3 (0.5) | 0 | 0.5 | 0-1(0.5) | 0.5-2(1) | 1-3 (2) |  |
| ATN, n (%) |  |  |  |  |  |  |  |
| A+T-N- | 9 (8.8%) | 5 (63%) | 4 (27%) | 0 (0%) | 0 (0%) | 0 (0%) |  |
| A+T-N+ | 1 (1.0%) | 0 (0%) | 0 (0%) | 0 (0%) | 1 (3.2%) | 0 (0%) |  |
| A+T+N- | 6 (5.9%) | 2 (25%) | 2 (13%) | 1 (2.9%) | 0 (0%) | 1 (7.1%) |  |
| A+T+N+ | 86 (84%) | 1 (13%) | 9 (60%) | 33 (97%) | 30 (97%) | 13 (93%) |  |

MMSE: Mini-mental State Examination, CDR: Clinical Dementia Rating, A: CSF amyloid β42, T: CSF phosphorylated Tau181, N: CSF total tau
<sup>1</sup>Kruskal-Wallis rank sum test; Fisher's exact test

### Diagnostic and differential diagnosis performance of plasma biomarkers

The mean NPQ (NULISA Protein Quantification) values of pTau217 showed significant differences between AD and the other groups, with a very large effect size (p<0.001; η²p = 0.45). DLB patients had 1.3 times higher pTau217 NPQ values than MCI patients (p = 0.03). The NPQ values of pTau181 and pTau231 between AD patients and other groups was also significantly different (p <0.001; η²p =0.30 with a 95% CI (0.22–1) and p<0.001; η²p =0.37 with a 95% CI (0.29–1), respectively). Like pTau217, pTau181 and pTau231 were significantly higher in DLB patients compared to MCI (p=0.003 and p=0.001 respectively). Also FTD patients’ mean NPQ values of pTau231 were slightly higher than MCI patients (p=0.04).

Plasma mean Aβ42 NPQ values was highest in the DLB group with a moderate effect size (p=0.002; η²p = 0.07 with a 95% CI (0.02–1), p values for DLB vs AD was 0.009, DLB vs MCI 0.008, DLB vs FTD 0.002).

The median NfL NPQ value was highest in FTD patients with relatively large effect size (p < 0.001; ε2 = 0.2 with a 95% CI (0.12–1)). AD and DLB group’s median NfL levels were higher than the MCI group’s (1.5 times difference with p<0.001 and 5 times difference with p <0.001, respectively).

The mean plasma GFAP NPQ value was highest in the AD group with a large effect size (p<0.001; η²p = 0.28 with a 95% CI (0.19–1). Also it was higher in the DLB group compared to the MCI and FTD groups (1.5 times difference with p=0.004 and 1.5 times difference with p=0.009 respectively)(**Figure 1).**

**Figure 1.**
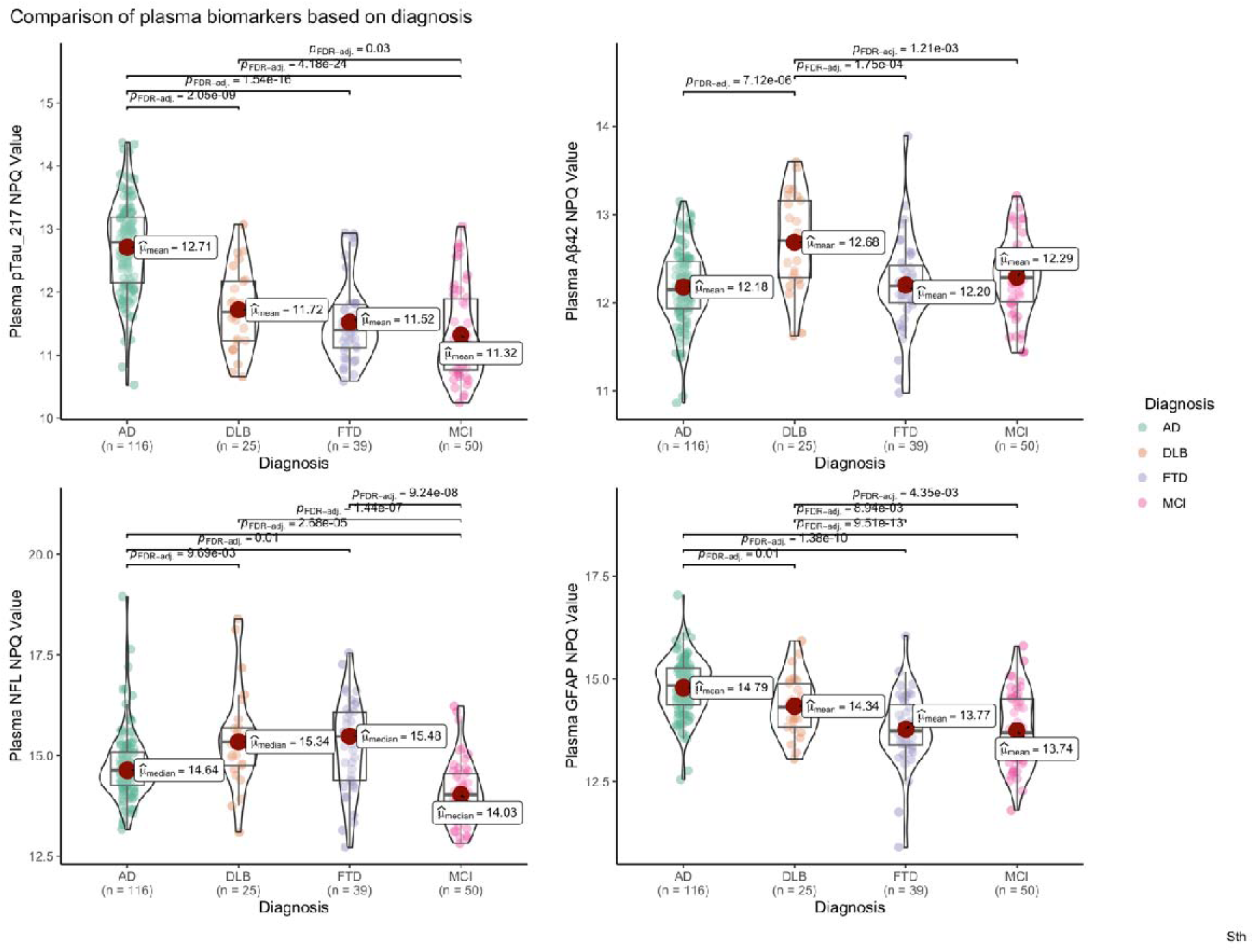
Violin plots illustrating plasma biomarker levels of pTau217, Aβ42, NfL, and GFAP across patients with AD, DLB, FTD, and MCI. Each violin plot shows the distribution of normalized NPQ values for each biomarker within the diagnostic groups, with mean or median values highlighted by red squares. Statistically significant differences between diagnostic groups were assessed using adjusted p-values (p<0.05, FDR-adj indicated) from group comparisons. NPQ: Normalized Protein Quantification, pTau-217: Phosphorylated tau-217, Aβ42: amyloid-beta 42, NfL: neurofilament light chain, GFAP: glial fibrillary acidic protein, AD: Alzheimer’s Disease, DLB: Dementia with Lewy Bodies, FTD: Frontotemporal Dementia, MCI: Mild Cognitive Impairment, FDR-adj: Post hoc pairwise Dunn’s test with FDR (False Discovery Rate) adjustment

We evaluated the ability of different plasma markers to detect amyloid pathological change (A+) defined by CSF and diagnosis. When assessing the potential of plasma pTau217 values to differentiate between A+ and A− status, AUC was 0.87 (95%CI 0.83–0.92) with 0.82 sensitivity and 0.83 specificity. Accuracy was 0.82 PPV (positive predictive value) 0.89 and NPV (negative predictive value) 0.72 (**Figure 2**).

**Figure 2.**
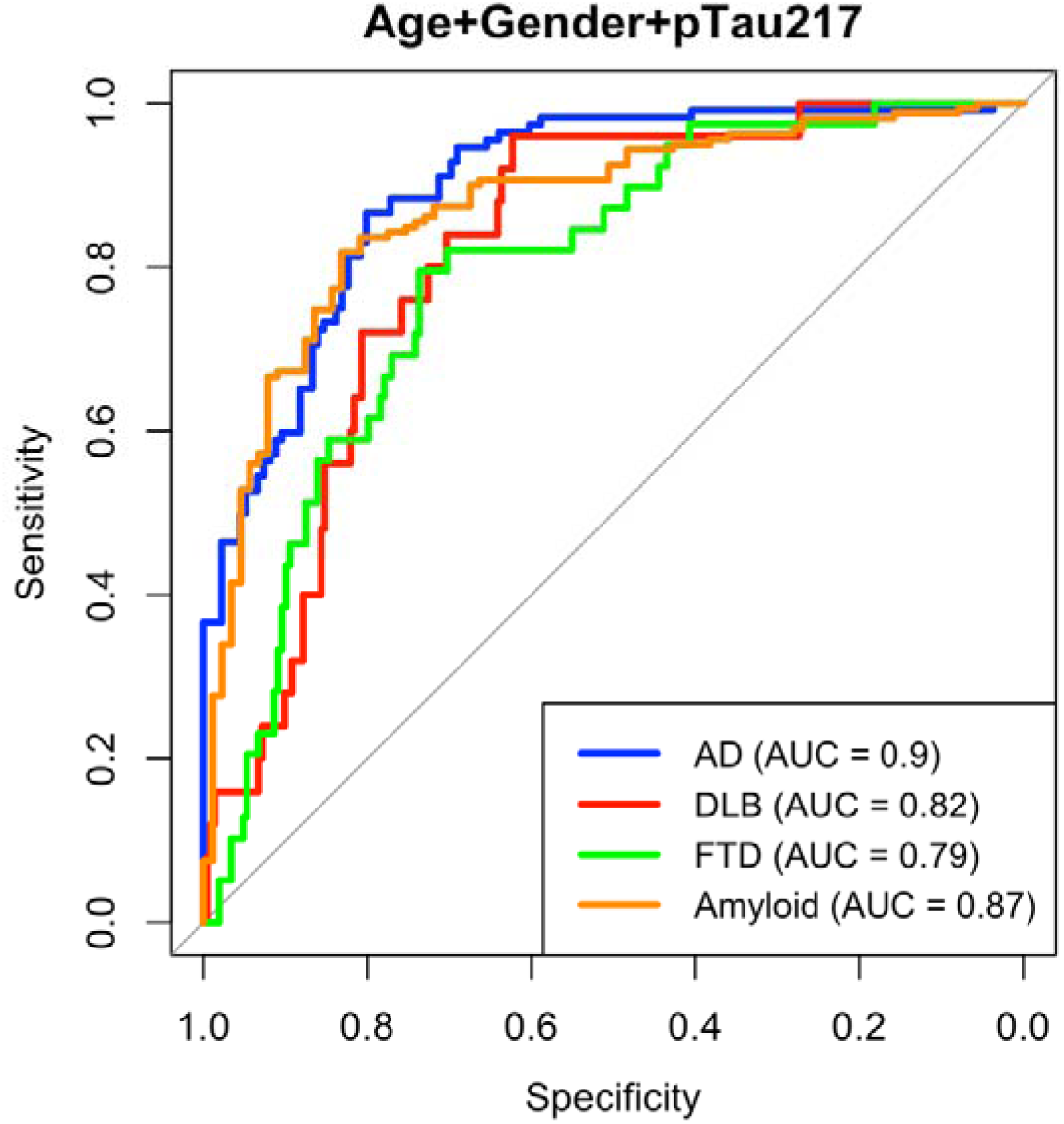
ROC curves for CSF amyloid status, AD, DLB and FTD diagnosis. The ROC curves display the diagnostic performance of a model combining plasma pTau217 levels, age, and gender for predicting AD, DLB, FTD diagnoses, and amyloid status in cerebrospinal fluid (CSF), separately. The AUC for AD, DLB, FTD, and amyloid positivity are 0.9, 0.82, 0.79, and 0.87, respectively. AD: Alzheimer’s Disease, DLB: Dementia with Lewy Bodies, FTD: Frontotemporal Dementia, AUC: Area Under the Curve

Plasma pTau217 value showed the best performance for predicting AD (AUC 0.9 [95%CI 0.86–0.94], 0.87 sensitivity and 0.8 specificity with accuracy 0.79, PPV 0.81 and NPV 0.78), DLB (AUC 0.84 [95%CI 0.76–0.91], 0.8 sensitivity and 0.8 specificity with accuracy 0.9, PPV 0.5 and NPV 0.9) and FTD diagnosis (AUC 0.79 [95%CI 0.72–0.86], 0.79 sensitivity and 0.74 specificity with accuracy 0.83, PPV 0 and NPV 0.84). ORs of these models are shown in **Supplementary table 1.**

Next, we studied how the combination of different markers predicted amyloid status. Adding GFAP to pTau217 increased the AUC to 0.89 (95%CI 0.84–0.93) but there were no differences between models (p=0.12). Also adding Aβ40, Aβ42 or Aβ42/40 data didn’t improve the model.

FABP3 (fatty acid-binding protein) had a similar performance as pTau217 for DLB diagnosis (AUC 0.83 [95%CI 0.76–0.90], 0.84 sensitivity and 0.75 specificity). When FABP3 was added pTau217 the AUC was 0.87 (95%CI 0.8–0.94) but this did not improve the model (p=0.08). Also GFAP in DLB showed a similar performance for diagnosis (AUC 0.82 [95%CI 0.75–0.89]).

When adding NfL data to pTau217 for FTD diagnosis the AUC increased to 0.88 (95% CI 0.83–0.92) with 0.97 sensitivity and 0.67 specificity p<0.01).

### Plasma and CSF biomarker correlation

In the overall cohort, CSF Aβ42 negatively correlated with plasma pTau, with a slight improvement for plasma pTau/Aβ42. CSF pTau181 and tTau positively correlated with plasma pTau, with a correction for CSF Aβ42 enhancing these relationships. MMSE moderately correlated with plasma pTau and NfL (**Figure 3**).

**Figure 3.**
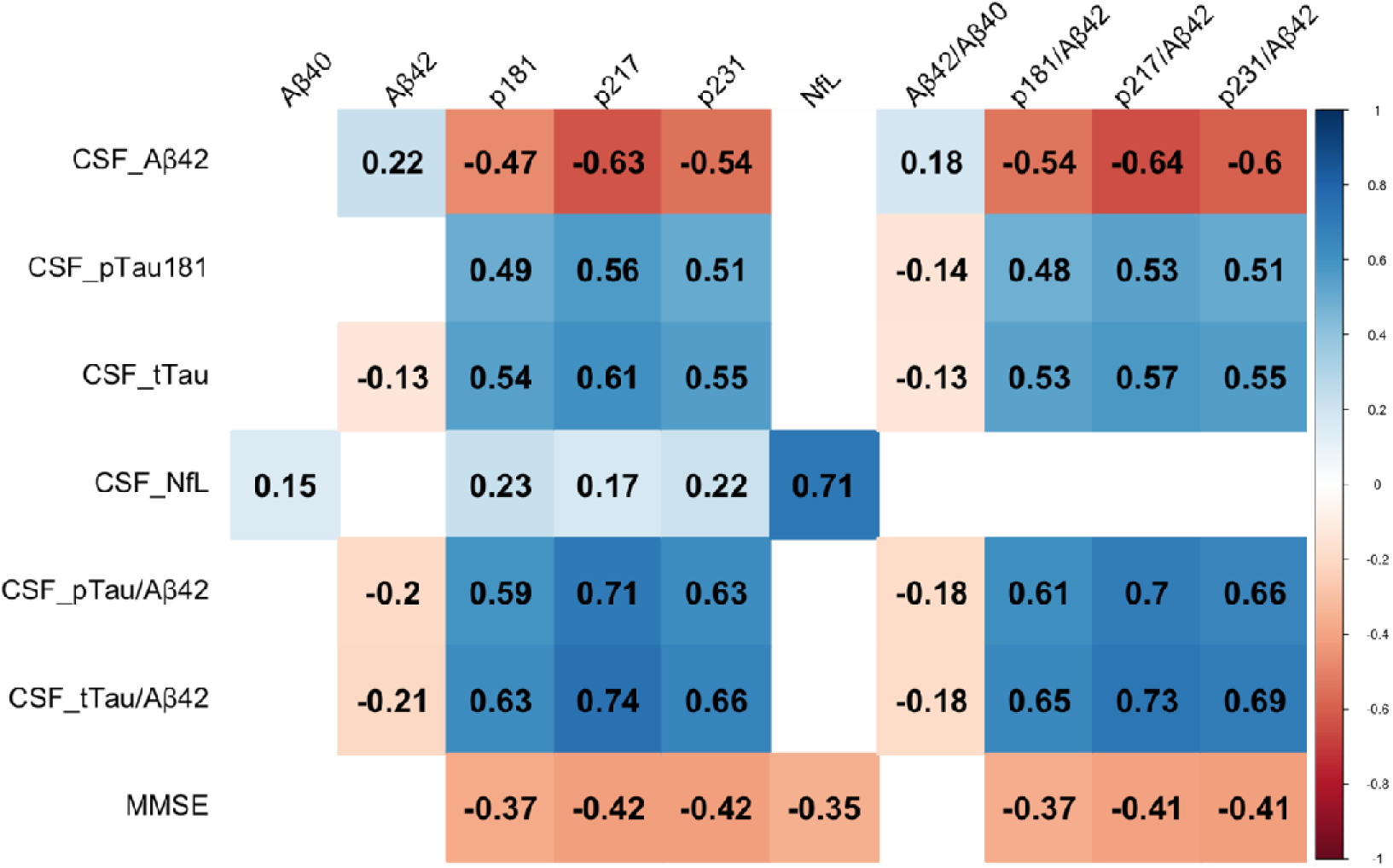
Correlation heatmap between CSF and plasma biomarkers for all cohort. The heatmap displays the correlation between CSF biomarkers and MMSE on the y-axis and plasma biomarkers on the x-axis. The numbers represent Spearman correlation coefficients, indicating the strength and direction of the relationships between the variables. Blue indicates positive correlations, and red indicates negative correlations. Only FDR corrected p value < 0.05 are showed. Aβ42: Amyloid-beta 42, Aβ40: Plasma Amyloid-beta 40, pTau181: Phosphorylated tau 181, tTau: Total tau, NfL: Neurofilament light chain, pTau/Aβ42: Ratio of phosphorylated tau 181 to Aβ42, tTau/Aβ42: Ratio of total tau to Aβ42, MMSE: Mini-Mental State Examination score, p181: Phosphorylated tau-181, p217: Phosphorylated tau-217, p231: Phosphorylated tau-231, Aβ42/Aβ40: Ratio of Aβ42 to Aβ40, p181/Aβ42: Ratio of phosphorylated tau 181 to Aβ42, p217/Aβ42: Ratio of phosphorylated tau 217 to Aβ42, p231/Aβ42: Ratio of phosphorylated tau 231 to Aβ42.

When analysed by diagnosis, CSF Aβ42 positively correlated with plasma Aβ42 in DLB and FTD, and with plasma Aβ40 only in DLB. In AD, CSF Aβ42 negatively correlated with plasma pTau217. Correcting plasma pTau for Aβ42 strengthened the correlations in FTD and DLB. The strongest CSF-plasma pTau correlations were observed in DLB, with a further improvement after Aβ42 correction. Notably, A+TxN-(CSF amyloid +, Tau + or -, and neurodegeneration -) MCI showed markedly increased CSF pTau181 and plasma pTau correlations. CSF pTau181 correlated with the Aβ42/Aβ40 ratio in DLB. CSF tTau correlated with plasma pTau in AD and DLB, with the strongest correlations in A+TxN-MCI for plasma pTau181. CSF NfL consistently correlated with plasma NfL across all groups **(Supplementary Fig. 1).**

All plasma pTau biomarkers correlated with CSF pTau181/Aβ42 in DLB and AD, with stronger correlations in DLB. While correlations for CSF and plasma biomarkers were low in AD patients, their trajectories through the AD continuum displayed a similar pattern **(Supplementary Fig. 2)**.

Regarding CSF AT status, CSF Aβ42 moderately correlated with plasma Aβ42/Aβ40 only in A-T+. It negatively correlated with plasma pTau biomarkers in A+T- and A-T+ groups, with no correlations in A-T- or A+T+. Correcting plasma pTau for Aβ42 improved A+T- and A-T+ correlations and revealed correlations in A+T+ group **(Supplementary Fig. 3).**

CSF Aβ42 negatively correlated with GFAP in DLB. CSF NFL correlated with plasma GFAP in AD, and negatively correlated with plasma GFAP/NfL in AD, DLB, and FTD. CSF NfL also correlated with plasma tTau and FABP3 in DLB, and with FABP3 in FTD. CSF tTau correlated with plasma total Tau and FABP3 in DLB, and with tTau and GFAP in A+TxN-MCI. MMSE was negatively correlated with all plasma pTau with the strongest correlation observed for pTau217 (r = -0.51, p < 0.01). Additionally, MMSE negatively correlated with plasma GFAP, tTau, and NfL in this group. In the DLB group MMSE is negatively correlated with plasma GFAP and tTau. And MMSE is negatively correlated with GFAP in the FTD group.

Plasma HTT, SNCA, SOD1 and TARDBP biomarkers were highly correlated with each other (HTT vs SNCA r= 0.89, HTT vs SOD1 r=0.87, HTT vs TARDBP r:0.93, SNCA vs SOD1 r: 0.9, SNCA vs TARDBP r:0.87, SOD1 vs TARDPB r: 0.83; all adjusted p values <0.001) **(Supplementary Fig. 4).** And all of them were highly correlated with FGF2, ARSA, ANXA5, IL18, MDH1, PGK1, RUVBL2 (r:0.64-0.92). These high correlations persisted or even increased, when subdividing different disease groups.

### Biomarker Differentiation Across Neurodegenerative Diseases and AD Stages

In comparison with the AD group, plasma pTau181, pTau217 and pTau231 had significantly lower odds for DLB (OR 0.21, 95% CI: 0.08–0.53, p= 0.005; OR 0.14, 95% CI: 0.05–0.4, p= 0.001 and OR 0.18, 95% CI: 0.07–0.47, p= 0.003, respectively) and for FTD group (OR 0.13, 95% CI: 0.05–0.32, p < 0.001; OR 0.08, 95% CI: 0.05–0.35, p < 0.001 and OR 0.14, 95% CI: 0.05–0.35, p < 0.001, respectively), controlling for age and gender (**Figure 4**). Total tau and GFAP also had showed lower odds for both DLB (OR 0.32 and 0.25) and FTD (OR 0.2 and 0.18) groups.

**Figure 4.**
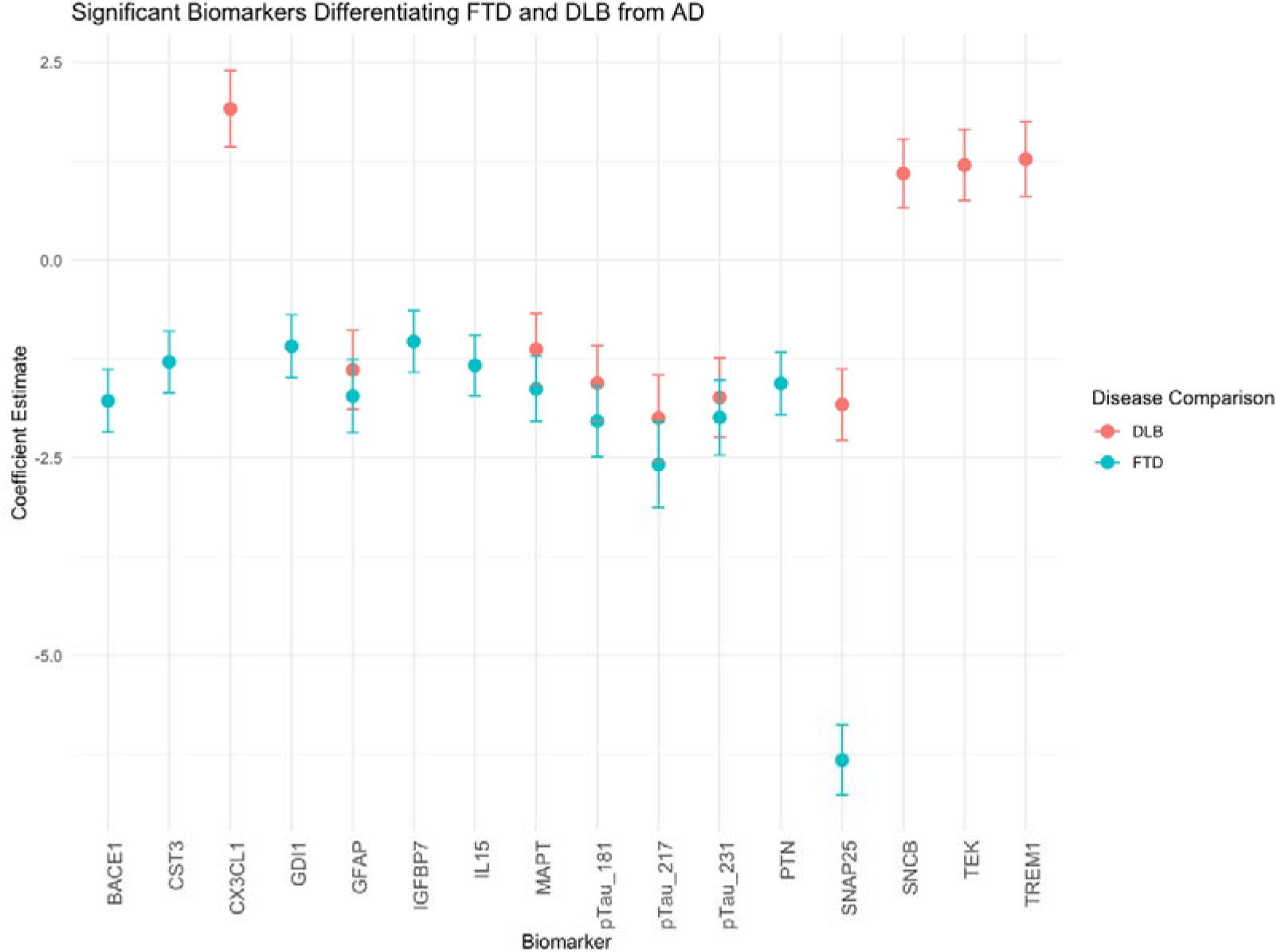
Multinomial Logistic Regression Coefficients for Biomarkers Differentiating DLB and FTD from AD. Biomarkers that above zero are increased in DLB compared to AD. Biomarkers that below zero are increased in AD compared to DLB or FTD. FTD: Frontotemporal Dementia, DLB: Dementia with Lewy Bodies, AD: Alzheimer’s Disease, BACE1: beta-site amyloid precursor protein cleaving enzyme 1, CST3: cystatin C, CX3CL1: chemokine (C-X3-C motif) ligand 1 (fractalkine), GDI1: GDP dissociation inhibitor 1, GFAP: glial fibrillary acidic protein, IGFBP7: insulin-like growth factor binding protein 7, IL15: interleukin-15, MAPT: microtubule-associated protein tau, pTau181: phosphorylated Tau-181, pTau217: Phosphorylated Tau-217, pTau231: Phosphorylated Tau-231, PTN: pleiotrophin, SNAP25: synaptosomal-associated protein 25, SNCB: beta synuclein, TEK: tyrosine kinase, TREM1: triggering receptor expressed on myeloid cells 1

In comparison with the AD group, SCNB, TEK and TREM1 had higher odds ratios for the DLB group. When comparing with AD; BACE1, CST3, GDI1, IGFBP7, IL15, PTN and SNAP25 had lower odds for the FTD group. The DLB group also had lower odds for SNAP25.

In the analysis of biomarkers across the AD continuum, PGF was significantly associated with disease stage, with an OR of 5.7 (95% CI:1.8-18, *p* = 0.002), indicating a 5.7-fold increase in odds for more advanced stages per unit increase in PGF levels. Similarly, pTau217 (OR=4.2, CI: 2.4-7.3, p<0.001), pTau181 (OR=4.2, CI:2.2-8.2, p<0.001), pTau231 (OR=4.1, CI: 2.3-7.5, p<0.001), SNAP25 (OR=3.9, CI:3.2-4.8, p<0.001), tTau (OR=2.9, CI:1.5-5.6, p=0.002), GFAP (OR=2.6, CI: 1.6-4.4, p<0.001) and NfL (OR=2.4, CI: 1.5-3.7, p <0.001) showed a significant association with disease progression (**Figure 5**).

**Figure 5.**
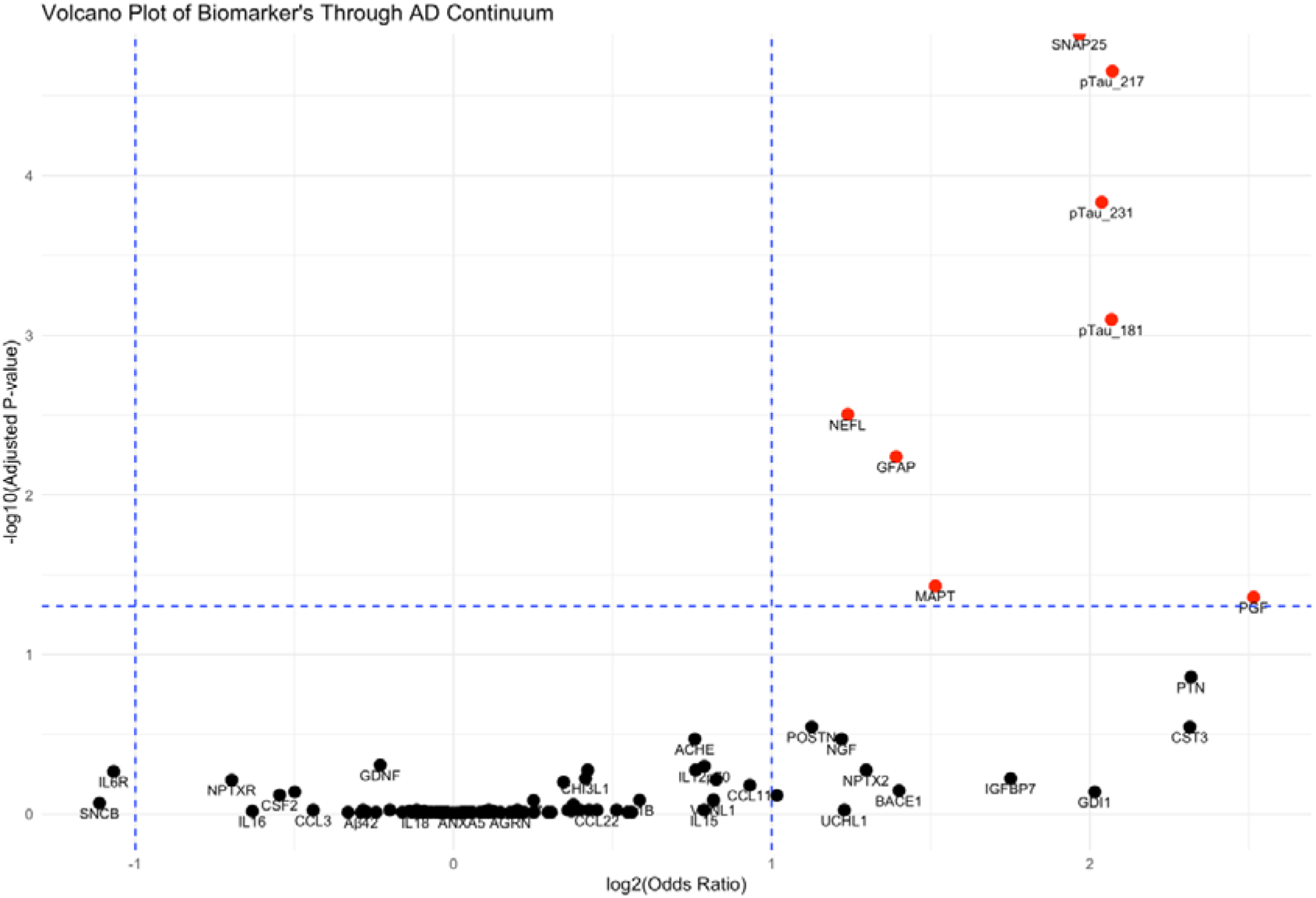
Volcano Plot of Biomarkers Across the AD Continuum Based on Ordinal Regression. Red biomarkers are significantly increasing with disease stage. AD: Alzheimer’s Disease, pTau181: phosphorylated tau-181, pTau217: phosphorylated tau-217, pTau231: phosphorylated tau-231, SNAP25: synaptosomal-associated protein 25, NEFL: neurofilament light chain, GFAP: glial fibrillary acidic protein, MAPT: microtubule-associated protein tau, PGF: placental growth factor

We extracted all plasma biomarkers with age and gender into the RF classification model and calculated feature importance. ROC analysis for the diagnosis of AD, FTD and DLB and the top 20 features are shown in **Figure 6**. Accuracy of this model was 0.73.

**Figure 6.**
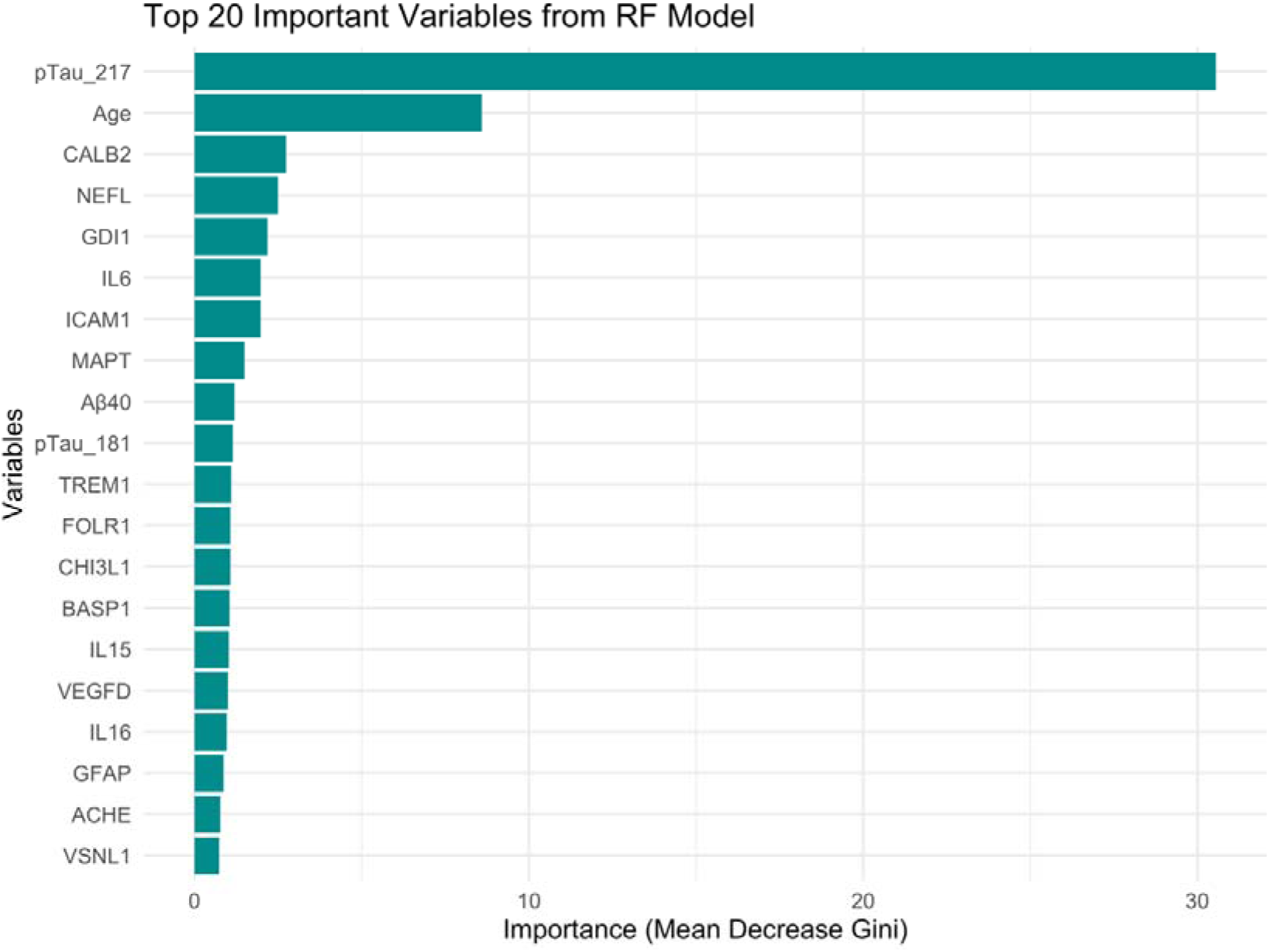
*Top 20 Important Variables for Differentiating AD, DLB, and FTD using a Random Forest Model*. The plot ranks the top 20 most important variables based on the Mean Decrease Gini score from the Random Forest model. pTau_217: phosphorylated tau-217, CALB2: calretinin, NEFL: neurofilament light chain, GDI1: GDP dissociation inhibitor 1, IL6: interleukin 6, ICAM1: intercellular adhesion molecule 1, MAPT: microtubule-associated protein tau, Aβ40: amyloid beta 40, pTau_181: phosphorylated tau-181, TREM1: triggering receptor expressed on myeloid cells 1, FOLR1: folate receptor 1, CHI3L1(YKL40): chitinase-3-like protein 1, BASP1: brain acid soluble protein 1, IL15: interleukin 15, VEGFD: vascular endothelial growth factor D, IL16: interleukin 16, GFAP: glial fibrillary acidic protein, ACHE: acetylcholinesterase, VSNL1: visinin like 1

ROC-AUC of all features for distinguishing AD from other groups was 0.85 [95% CI: 0.81–0.88], FTD from other groups was 0.83 [95% CI: 0.79–0.88] and DLB from other groups was 0.75 [95% CI: 0.69–0.82] (**Supplementary Fig. 5**). The first 10 important biomarkers for AD, FTD and DLB are shown in **Supplementary Fig. 6-8**.

When we took the biomarkers that we find significant in multinomial logistic regression model our random forest model improved for all categories. AUC for AD was 0.88 [95% CI: 0.84–0.91], FTD was 0.87 [95% CI: 0.83–0.91], and DLB was 0.81 [95% CI: 0.76–0.86]. Accuracy for whole model was 0.76. (**Supplementary Fig. 9-13**).

### Gene Ontology Pathway Enrichment

GO analysis of differentially measured biomarkers across AD, DLB, and FTD revealed a significant enrichment in biological processes related to cell and neuron projection development, modulation of chemical synaptic transmission, and regulation of trans-synaptic signalling. Cellular components primarily involved in these processes included the neuronal cell body, endoplasmic reticulum lumen, and glial cell projection **(Supplementary Fig. 14).** In terms of molecular functions integrin binding was a significant function associated with GFAP, PTN and CX3CL1.

## Discussion

In the present study, we investigated plasma biomarkers related to different aspects of neurodegeneration. We assessed their diagnostic accuracy, correlation with established CSF biomarkers, and conducted exploratory analyses within a mixed cohort of patients with MCI, AD, DLB, FTD, and other dementias. We found that plasma pTau217 has the highest diagnostic accuracy for AD, DLB and FTD diagnosis as well as CSF amyloid status. Also we found novel plasma proteomic alterations for different dementia types and therefore promising biomarker candidates for future research.

Consistent with the previous literature, all three plasma pTau forms and GFAP exhibited the highest normalized protein expression quantification (NPQ) values in the AD group, while Aβ42 was highest in the DLB group ^37^. Across the entire cohort, plasma pTau217, pTau181, and pTau231 showed strong correlations with CSF Aβ42, pTau181, and tTau. These correlations were further enhanced when pTau values were adjusted for Aβ42 in both CSF and plasma. Additionally, CSF and plasma NfL demonstrated a high degree of correlation.

In our mixed dementia cohort, plasma pTau217 demonstrated the best performance for diagnosing AD, DLB, and FTD, as well as for predicting CSF amyloid positivity (AUC 0.9, 0.84, 0.79, and 0.87, respectively). This finding aligns with a recent study employing thirty-three different p-tau biomarker assays, which reported superior fold-changes, discriminative accuracies for detecting AD pathology, and inter-platform agreement for plasma pTau217 measurements compared to other plasma pTau variants ^38^. Plasma pTau217, evaluated on the NULISA platform, achieved an AUC of 0.91 for amyloid PET positivity ^39^. These recent studies collectively highlight plasma pTau biomarkers, particularly pTau217, as the most sensitive plasma biomarkers for AD diagnosis, even in its early stages ^40^.

The observed association between disease progression and pTau217, pTau181, pTau231, tTau, NfL, and GFAP aligns with the literature ^41–44^. Our findings also support recent studies indicating the involvement of another neurodegeneration biomarker SNAP25 ^45^ and PGF, a VEGF family member, in disease progression ^46^. Within our cohort, the addition of NfL to pTau217 significantly improved the ROC AUC for FTD (from 0.79 to 0.88), similar to previous reports ^43,47^. Although plasma GFAP and FABP3 levels exhibited comparable performance to pTau217, their inclusion to the model did not significantly enhance AUC ^48–50^.

In line with previous research, plasma pTau and NfL showed a negative correlation with MMSE scores specifically within the AD group ^41^. Additionally, plasma GFAP levels were negatively correlated with MMSE scores across AD, FTD, and DLB groups ^51,52^. The predictive value of CSF and plasma pTau for future cognitive decline has yielded conflicting results in previous studies ^41,42,53^, likely due to variations in study populations. To establish a reliable cutoff for predicting cognitive decline, longitudinal studies are necessary.

In the DLB group, GFAP and pTau levels were elevated compared to the MCI group, and CSF Aβ42 was negatively correlated with GFAP. Furthermore, all three plasma pTau forms correlated with CSF pTau181, and pTau217 and this effectively discriminated the DLB group from other diagnostic categories.

These results align with a study that evaluated amyloid and tau PET imaging alongside plasma biomarkers in the DLB continuum ^48^. This study reported the highest accuracy of plasma pTau181 in detecting abnormal amyloid PET in DLB patients. Although the study did not directly examine correlations with Aβ42, it did not find any significant associations between amyloid PET and plasma Aβ42/Aβ40 ratios. Similarly, another study found no differences in plasma Aβ42/Aβ40 levels between PET-Aβ negative and PET-Aβ-positive DLB cases ^54^. In contrast to the AD group, our analysis revealed a correlation between CSF Aβ42 and plasma Aβ40 and Aβ42 in the DLB group.

When stratified by diagnosis, correlations between plasma pTau and CSF biomarkers diminished in AD, with no significant correlation with CSF pTau. Plasma pTau217 exhibited a slight negative correlation with CSF Aβ42 (r=-0.21), and plasma pTau217, pTau181, and pTau231 showed weak positive correlations with CSF tTau. Notably, no significant correlations were observed with CSF pTau.

This data aligns with prior research showing weak correlations between plasma and CSF pTau in AD ^55,56^, and a dissociation between tau PET and CSF pTau in AD patients ^44,53^. A recent study that employing thirty-three different pTau biomarker assays found no correlations between plasma and CSF pTau levels within the AD group ^38^. These findings suggest fluid and imaging biomarkers capture distinct aspects of tau pathophysiology, reflected in the updated ATN classification (T1: CSF/plasma, T2: tau PET/oligomeric Tau) ^15^.

Imaging studies suggest soluble pTau is a better reflection of Aβ42 plaque concentration than PET-measured tau ^57^. CSF tau levels are elevated even before symptom onset in individuals with Aβ+ pathology, coinciding with amyloid deposition and preceding PHF-tau detection by PET. This suggests that CSF tau increases in response to Aβ pathology, even without significant PHF-tau accumulation ^58^. This relationship is further supported by studies showing correlation between increased CSF tau production and brain amyloid levels ^59^, and by the absence of increased CSF pTau in individuals with MAPT mutations and tau pathology but no amyloid ^58^. Additionally, the presence of tau within amyloid plaques suggests that initial tau deposits occur near Aβ plaques ^60–63^. Animal models, exhibiting Aβ plaques but not NFTs, show elevated CSF tau, further support the link between Aβ pathology and CSF tau increases ^64,65^. While tau PET imaging strongly correlates with neurofibrillary tangles ^66^, CSF tau biomarkers show a weaker correlation with postmortem tau aggregates ^67,68^.

Our findings, consistent with others, demonstrate the strongest CSF/plasma pTau correlation in amyloid-positive MCI ^16,58^, and a weakening of this correlation in later AD stages. This aligns with the observation that pTau-NFT correlation weakens in AD dementia and late Braak stages ^69,70^.

CSF and plasma biomarkers in AD exhibit similar trajectories, with pTau and NfL levels initially increasing with disease severity, then plateauing and declining (**Figure 3).** This pattern reflects the early increase and subsequent decrease of CSF pTau in symptomatic AD ^69,71–73^. Soluble pTau peaks during maximal amyloid load and declines, likely due to sequestration within NFTs ^74–76^, mirroring animal models where CSF pTau tracks Aβ deposition ^65^. However, tau PET signal may continue to increase ^77^, likely driven by oligomeric and conformational tau, as pTau staining plateaus but conformational and oligomeric tau staining increases ^78^. The early co-localization of soluble tau and Aβ pathology suggests a close relationship preceding mature NFT emergence ^78,79^.

Based on the evidence presented and existing literature, Tau accumulation initiates around amyloid plaques, with tau release and phosphorylation triggered by Aβ deposition. CSF amyloid and plasma pTau levels correlate in amyloid+ MCI, but this weakens in AD, suggesting pTau reflects amyloid pathology early on. As PHF formation progresses, pTau correlation weakens despite concentration increases, implying pTau, initially sequestered as dystrophic neurites, later becomes a substrate for PHF formation, explaining the nonlinear variation of pTau levels across AD stages. This aligns with our DLB findings, where pTau maintains its amyloid correlation, unlike in AD. Although both AD and DLB have amyloid plaques, their composition differs, with DLB plaques containing minimal Aβ40 and showing a broader reduction in amyloid-beta species ^63,80,81^.

In our study, CSF pTau correlated with plasma pTau only in A+T-individuals, particularly in A+T-MCI patients, consistent with studies showing significance in A+ MCI ^82^ and cognitively normal A+ individuals ^83^.

We also found a correlation between CSF pTau/Aβ42 ratio and plasma pTau, even without a direct CSF-plasma pTau correlation in AD. The pTau/Aβ42 ratio enhanced correlations with DLB, FTD, and amyloid-positive MCI, consistent with its utility in discriminating AD and predicting amyloid PET positivity ^84–87^. These findings suggest pTau levels accompany amyloid pathology, with the pTau/Aβ42 ratio offering improved correlation and diagnostic potential.

While we found high correlations for HTT (huntingtin), SNCA (α-synuclein), SOD1 (superoxide dismutase-1), TARDBP (TAR DNA-binding protein 43), and other biomarkers, these are not typically correlated with CSF and have non-brain sources ^88–91^. Further investigation is needed to determine whether these correlations are due to peripheral origins, shared enzymatic alterations, or truly informative significant associations.

AGRN (agrin), CX3CL1 (fractalkine), SNCB (β-synuclein), TEK (tyrosine kinase), and TREM1 (Triggering receptor expressed on myeloid cells-1) are elevated in DLB patients compared to AD. TEK inhibitors decrease CSF a-synuclein and clinical trials are currently investigating TEK inhibitors for DLB treatment ^92^. TREM1 is elevated in CSF of individuals with AD compared to those with MCI. However, no significant difference in CSF TREM1 levels has been observed between AD and DLB ^93^. To date, there is a lack of sufficient research examining and comparing TREM1 levels in the plasma of AD and DLB patients.

CX3CL1, a chemokine primarily expressed by neurons, plays a crucial role in modulating microglial activity and mediating neuron-microglia communication. Pre-clinical studies have demonstrated the dual effects of fractalkine signalling on PD-related inflammation and degeneration. While clinical evidence remains limited, fractalkine may be implicated in PD progression ^94^. However, research on fractalkine’s role in DLB patients is not yet sufficient to draw conclusions.

CSF β-synuclein levels were found to be elevated only in patients with AD, but not in those with PD or DLB ^95^. The absence of β-synuclein expression in blood cells highlights its potential as a peripheral blood biomarker ^96^. Further investigation is needed due to the limited research in this area.

Multiple studies have detected agrin in AD plaques, cerebrovascular Aβ deposits, and neurofibrillary tangles ^61^. While in vitro studies and animal models have shown conflicting results regarding the potential impact of agrin on Aβ accumulation, its influence on brain Aβ deposition is evident ^97^. Agrin also induces conformational changes in α-synuclein, enhances its insolubility, and accelerates protofibril formation. Additionally, it colocalizes with α-synuclein in neuronal Lewy bodies within the substantia nigra of PD brains ^98^. To our knowledge, this study is the first to report elevated plasma agrin levels in DLB compared to AD.

Also we found elevated levels of BACE1 (beta-secretase-1), CST3 (cystatin C), GDI1 (GDP dissociation inhibitor-1), GFAP, IGFBP7 (insulin-like GF-binding protein-7), IL15, tTau, PGF (Placental Growth Factor), pTau, PTN (pleiotrophin), SLIT2 (slit guidance ligand 2), and SNAP25 (synaptosomal-associated protein-25) in AD patients compared to FTD patients.

GFAP, tTau, and all three pTau biomarkers were also increased in AD compared to DLB, with no differences observed between DLB and FTD patients for these biomarkers. These results align with previously discussed literature. BACE1, CST3, GDI1, IGFBP7, and IL15 are biomarkers associated with amyloid pathology and AD ^99–103^.

PGF levels in CSF have been found to be higher in FTD than in AD ^104^, but no plasma comparisons are available in literature. PTN is elevated in the CSF of AD patients compared to both cognitively normal individuals and those with other neurodegenerative disorders, and it correlates with tTau, pTau, and amyloid ^105^. Also in our GO analysis PTN, CX3CL1 and GFAP were found to be related to integrin binding. PTN is known for its role in neuronal growth and survival, CX3CL1 for mediating neuron-microglia communication. GFAP is a marker of astrocyte activation suggesting that integrin-mediated pathways could play a role in the progression and differentiation of these diseases, particularly through cellular adhesion, signalling mechanisms and, neuroinflammation. This finding aligns with previous research indicating that integrin pathways may be potential therapeutic targets in neurodegeneration ^106^.

SLIT2 was first identified as an axonal driver in the development of the CNS and appears to be involved in amyloid pathology, inflammation and blood-brain barrier integrity ^107,108^. SNAP25 is elevated in the CSF of AD patients compared to DLB and FTD ^109^, and in plasma compared to cognitively unimpaired individuals ^45^. To our knowledge, no studies have compared plasma SNAP25 levels between AD and other dementia types.

These findings align with our GO analysis, which revealed that differentially expressed biomarkers between diseases were involved in processes such as the regulation of neuron projection development, modulation and regulation of synaptic transmission, and trans-synaptic signalling.

Random forest analysis reveals that the overall importance is primarily driven by pTau and amyloid-related biomarkers. Among the top 20 important biomarkers, pTau217, NfL, tTau, pTau181, Aβ40, GFAP, and Aβ42 have been previously discussed. CALB2 (calretinin) has been identified as a hub gene in AD pathogenesis ^110^. UCHL1 has been reported to play a crucial role in PD and DLB, accelerating α-synuclein aggregation ^111^. Previous research has demonstrated decreased CSF levels and increased plasma levels of UCHL1 in PD patients, as well as reduced UCHL1 expression in the substantia nigra of DLB patients ^111,112^. To our knowledge, this study is the first to report elevated UCHL1 levels in DLB compared to AD. Furthermore, within our cohort, UCHL1 levels remained elevated in DLB patients relative to MCI patients. GDI1 and FABP3 are associated with microglial activation and are found to be increased in AD CSF and can differentiate between AD and genetic FTD patients ^113^. In response to decreased FOLR1 in CSF, brain folate supply to the cerebral cortex shifts from normal astrocytes to GFAP-positive astrocytes ^114^.

ICAM1 (CD200) is negatively correlated with neurofibrillary tangle, pTau, and amyloid plaque density in the brain tissue of DLB patients ^115^. A study measuring CSF proteomes to explore their association with known AD pathology found that YWHAZ (14-3-3 zeta/delta), SMOC1 (SPARC-related modular calcium-binding protein-1), FABP, CHI3L1, and BASP1 (brain acid soluble protein-1), all identified in our RF analysis, are related to amyloid or pTau ^116^.

VEGFD, associated with oligodendrocytes, increases with β-amyloid load and, unlike other VEGF receptors, is primarily involved in lymphangiogenesis ^117^. SMOC1 is elevated in the AD brain. Studies of autosomal dominant AD patients have shown that SMOC1 levels begin to rise 30 years before symptom onset ^118^ and correlate with NFT and amyloid pathology in numerous studies ^119^. However, no differences in SMOC1 levels were found between CU and AD serum samples. ANXA5 (annexin A5) is associated with familial late-onset AD ^120^ and exhibits higher plasma levels in AD and DLB cases compared to controls ^121^.

When evaluating synaptic dysfunction biomarkers in CSF, it was demonstrated that SNAP25, 14-3-3 zeta/delta, and NPTX2 (neuronal pentraxin), in various combinations, can discriminate AD from other neurodegenerative diseases ^122–124^. IL33 appears to be linked to tau pathology and microglial activation ^124^ and is closely associated with FTD ^125^. CHI3L1 (Chitinase-3-like protein-1 or YKL-40) is increased in the brain of sporadic FTD, while UCHL1 is increased in CSF, and NPTX1 and 2 are decreased in CSF in both sporadic and genetic FTD ^126^. PRDX6 (peroxiredoxin-6), an enzymatic protein unique to astrocytes in the CNS, regulates glial toxicity in tau-mediated neurodegeneration in AD and dopaminergic neurodegeneration in PD ^1,127^. CALB2 and other calcium-binding protein-expressing cortical neurons are not affected in DLB ^128^. MSLN (mesothelin), a tumor-associated antigen overexpressed on various malignant tumor cells, shows decreased mRNA levels in DLB patients compared to CU ^129^.

Despite the growing body of plasma proteomics research, few studies have examined the AD spectrum or compared AD to other common dementia types (FTD and DLB). This limits our understanding of AD-specific biomarkers and hinders progress toward differential diagnosis. Our findings suggest that there are distinct and diverse biological processes that are dysregulated in the different dementia types.

### Limitations

Our study is limited by the absence of data from healthy controls, which is crucial for assessing the utility of this technique in differentiating normal and pathological states and monitoring disease progression. But the patients were consecutively recruited, and the cohort composition reflects the characteristics commonly observed in memory clinic practice. Future longitudinal community-based studies should include both asymptomatic and symptomatic individuals to evaluate the real-world impact of plasma markers in clinical practice. Additionally, positive and negative predictive values should be assessed, considering the prevalence of AD pathology across age groups and the general population, to determine the screening potential of these markers.

The presence of positive AD CSF biomarker profile in 20–30% of patients clinically diagnosed with non-AD dementias raises the possibility of co-pathologies ^130^, another study limitation. Furthermore, the relatively young age of our AD group limits understanding of plasma pTau performance in older populations.

## Conclusion

The sensitivity and specificity of Tau PET, CSF and plasma pTau to identify abnormalities in tau metabolism differ. Integrating plasma biomarkers into clinical practice could revolutionize neurodegenerative disease diagnostics, improving accuracy, tracking progression, and monitoring treatment response. They may also uncover shared or specific pathways underlying different dementia types. However, further research is needed for validation in diverse cohorts and standardization for clinical use.

## Supporting information

Supplementary Fig. 1. CSF and plasma biomarkers correlation heatmap, based on diagnosis.

## Data availability

The data from this study will be made available in de-identified format to researchers upon request.

## Funding

The study is supported by the Scientific and Technological Research Council of Turkey-TUBITAK (Project No. 22AG017 – APYOK2. Grant Recipient: Erdinc Dursun). HZ is a Wallenberg Scholar and a Distinguished Professor at the Swedish Research Council supported by grants from the Swedish Research Council (#2023-00356, #2022-01018 and #2019-02397), the European Union’s Horizon Europe research and innovation programme under grant agreement No 101053962, Swedish State Support for Clinical Research (#ALFGBG-71320), the Alzheimer Drug Discovery Foundation (ADDF), USA (#201809-2016862), the AD Strategic Fund and the Alzheimer’s Association (#ADSF-21-831376-C, #ADSF-21-831381-C, #ADSF-21-831377-C, and #ADSF-24-1284328-C), the European Partnership on Metrology, co-financed from the European Union’s Horizon Europe Research and Innovation Programme and by the Participating States (NEuroBioStand, #22HLT07), the Bluefield Project, Cure Alzheimer’s Fund, the Olav Thon Foundation, the Erling-Persson Family Foundation, Familjen Rönströms Stiftelse, Stiftelsen för Gamla Tjänarinnor, Hjärnfonden, Sweden (#FO2022-0270), the European Union’s Horizon 2020 research and innovation programme under the Marie Skłodowska-Curie grant agreement No 860197 (MIRIADE), the European Union Joint Programme – Neurodegenerative Disease Research (JPND2021-00694), the National Institute for Health and Care Research University College London Hospitals Biomedical Research Centre, and the UK Dementia Research Institute at UCL (UKDRI-1003). J.H., D.A.S., and U.Y, are supported by the UK Dementia Research Institute [award numbers UK DRI-1009] through UK DRI Ltd, principally funded by the Medical Research Council. J.H., M.S., and D.A.S. are supported by the Dolby Foundation, and by the National Institute for Health Research University College London Hospitals Biomedical Research Centre. J.H. also received funding from Cure Alzheimer’s Fund.

## Conflicts of interest

HZ has served at scientific advisory boards and/or as a consultant for Abbvie, Acumen, Alector, Alzinova, ALZpath, Amylyx, Annexon, Apellis, Artery Therapeutics, AZTherapies, Cognito Therapeutics, CogRx, Denali, Eisai, LabCorp, Merry Life, Nervgen, Novo Nordisk, Optoceutics, Passage Bio, Pinteon Therapeutics, Prothena, Quanterix, Red Abbey Labs, reMYND, Roche, Samumed, Siemens Healthineers, Triplet Therapeutics, and Wave, has given lectures sponsored by Alzecure, BioArctic, Biogen, Cellectricon, Fujirebio, Lilly, Novo Nordisk, Roche, and WebMD, and is a co-founder of Brain Biomarker Solutions in Gothenburg AB (BBS), which is a part of the GU Ventures Incubator Program (outside submitted work). AJH has consulted for Quanterix and Lilly.

## Notes

### Author Declarations

Istanbul University Ethics Committee (approval number 2023/1191, granted 22/06/2023).

