## Supplementary Fig. 1. CSF and plasma biomarkers correlation heatmap, based on diagnosis. for "Exploratory Blood Biomarker Patterns in a Mixed Dementia Cohort"

***Supplementary Table 1.*** *ROC Models*

|  | CSF Amyloid positivity | | | AD Diagnosis | | | FTD Diagnosis | | | | DLB Diagnosis | | |
| --- | --- | --- | --- | --- | --- | --- | --- | --- | --- | --- | --- | --- | --- |
| **Characteristic** | **OR**^1^ | **95% CI**^1^ | **p-value** | **OR**^1^ | **95% CI**^1^ | **p-value** | **OR**^1^ | **95% CI**^1^ | **p-value** | **OR**^1^ | | **95% CI**^1^ | **p-value** |
| Age | 1.03 | 0.99, 1.07 | 0.2 | 0.99 | 0.95, 1.03 | 0.6 | 1.18 | 1.10, 1.28 | *<0.001* | 0.93 | | 0.89, 0.97 | *<0.001* |
| Gender | 0.78 | 0.40, 1.53 | 0.5 | 1.11 | 0.55, 2.27 | 0.8 | 1.72 | 0.69, 4.48 | 0.2 | 0.65 | | 0.30, 1.37 | 0.3 |
| pTau_217 | 7.87 | 4.86, 13.7 | *<0.001* | 11.0 | 6.49, 20.1 | *<0.001* | 0.48 | 0.25, 0.85 | *0.011* | 0.42 | | 0.26, 0.63 | *<0.001* |
| AD: Alzheimer dementia, MCI: Mild cognitive impairment, FTD: Frontotemporal dementia, DLB: Dementia with Lewy bodies  ^1^OR = Odds Ratio, CI = Confidence Interval | | | | | | | | | | | | | |

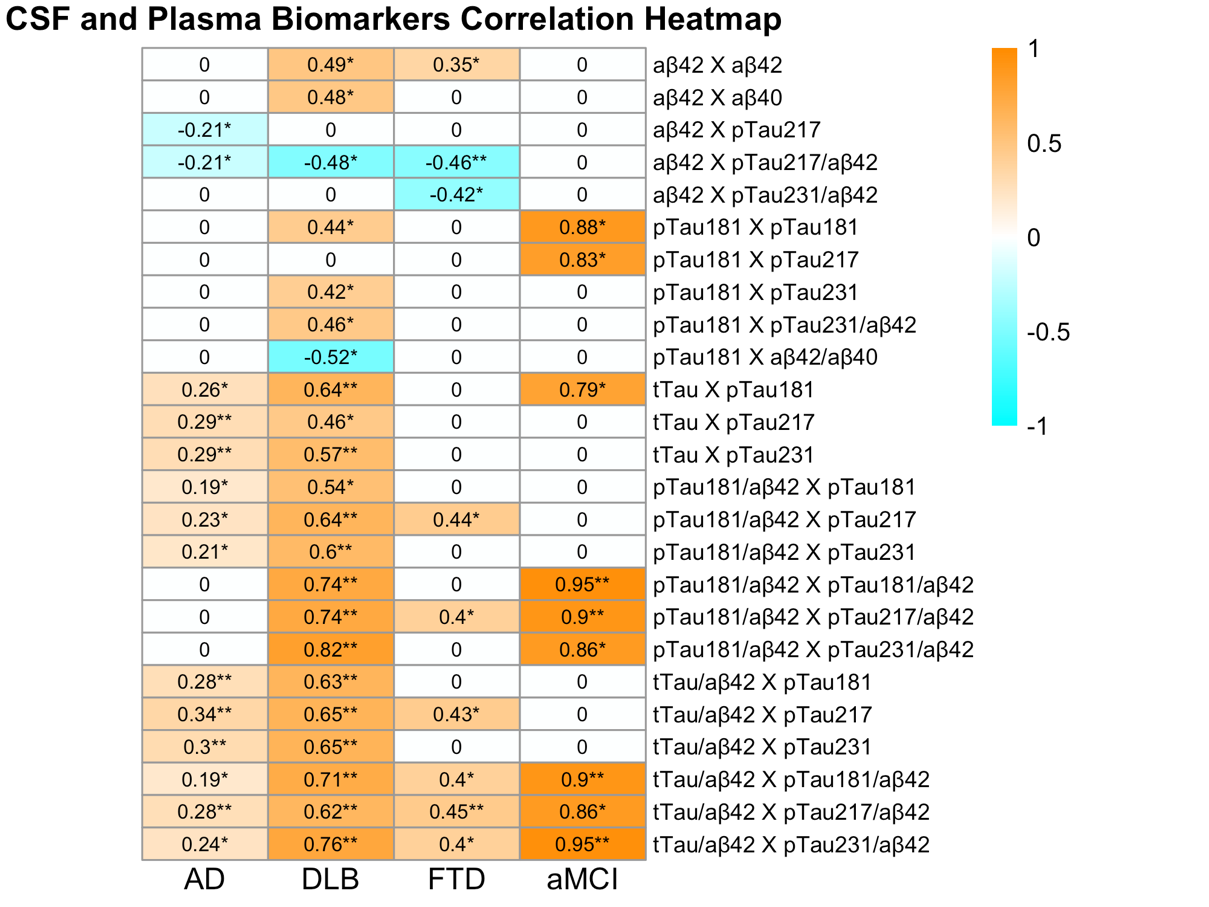
 ***Supplementary Fig.*** ***1.*** CSF and plasma biomarkers correlation heatmap, based on diagnosis. The heatmap represents the correlation between CSF and plasma biomarkers across diagnostic groups. First biomarker represents CSF and second biomarker represents plasma. The color scale represents correlation coefficients, with orange indicating positive correlations and blue indicating negative correlations. AD: Alzheimer’s Disease, DLB: Dementia with Lewy Bodies, FTD: Frontotemporal Dementia, aMCI: amyloid positive Mild Cognitive Impairment, Aβ42: amyloid beta 42, Aβ40: amyloid beta 40, pTau181: phosphorylated Tau-181, pTau217: phosphorylated tau-217, pTau231: phosphorylated tau-231, tTau: total tau. * p<0.05, ** p<0.01

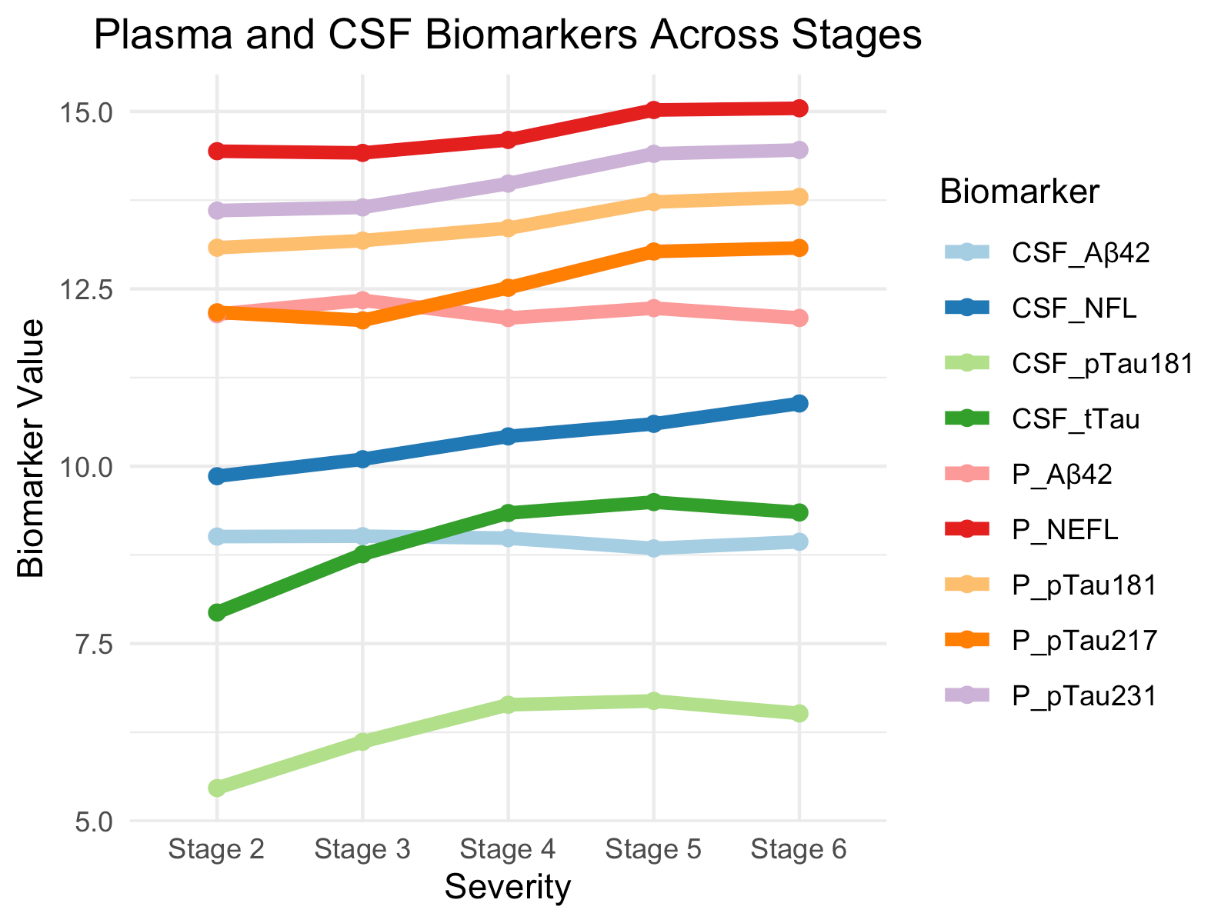

***Supplementary Fig.*** ***2.*** Plasma and CSF biomarkers’ trajectory across AD continuum. This figure displays the changes in various plasma and CSF biomarkers across AD stages of disease severity. The lines represent the mean biomarker values as the disease progresses, allowing for comparisons between plasma and CSF biomarkers at different stages. CSF_Aβ42: Cerebrospinal Fluid amyloid beta 42, CSF_NFL: Cerebrospinal Fluid neurofilament light chain, CSF_pTau181: Cerebrospinal Fluid phosphorylated tau-181, CSF_tTau: Cerebrospinal Fluid total tau, P_Aβ42: Plasma amyloid beta 42, P_NEFL: Plasma neurofilament light chain, P_pTau181: Plasma phosphorylated tau-181, P_pTau217: Plasma phosphorylated tau-217, P_pTau231: Plasma phosphorylated tau-231

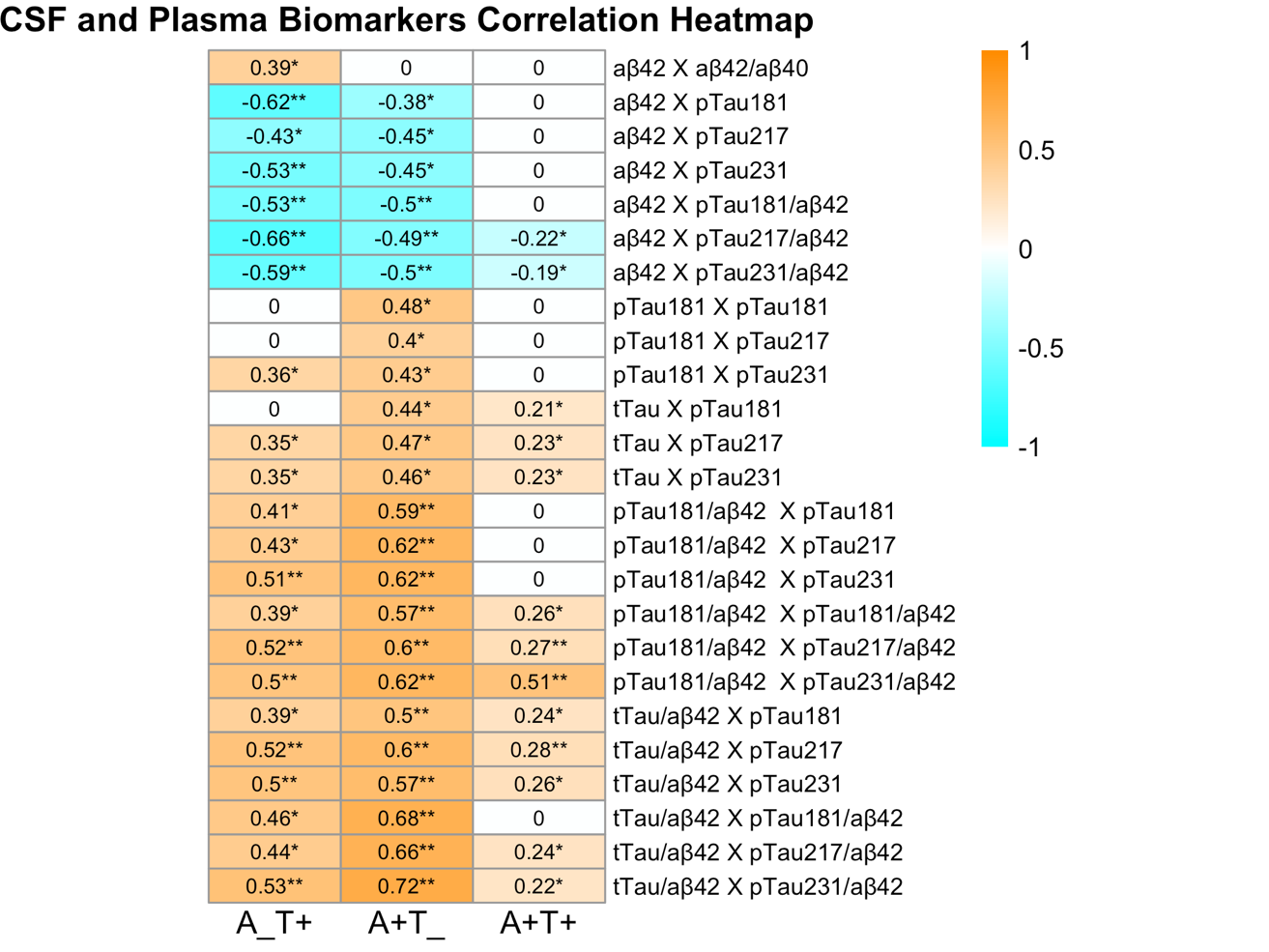

***Supplementary Fig.*** ***3.***CSF and plasma biomarkers correlation heatmap, based on CSF AT status***.*** The heatmap represents the correlation between CSF and plasma biomarkers for different CSF AT status. First biomarker represents CSF and second biomarker represents plasma. The colour scale represents correlation coefficients, with orange indicating positive correlations and blue indicating negative correlations. Aβ42: amyloid beta 42, Aβ40: amyloid beta 40, pTau181: phosphorylated tau-181, pTau217: phosphorylated tau-217, pTau231: phosphorylated tau-231, tTau: total tau. * p<0.05, ** p<0.01

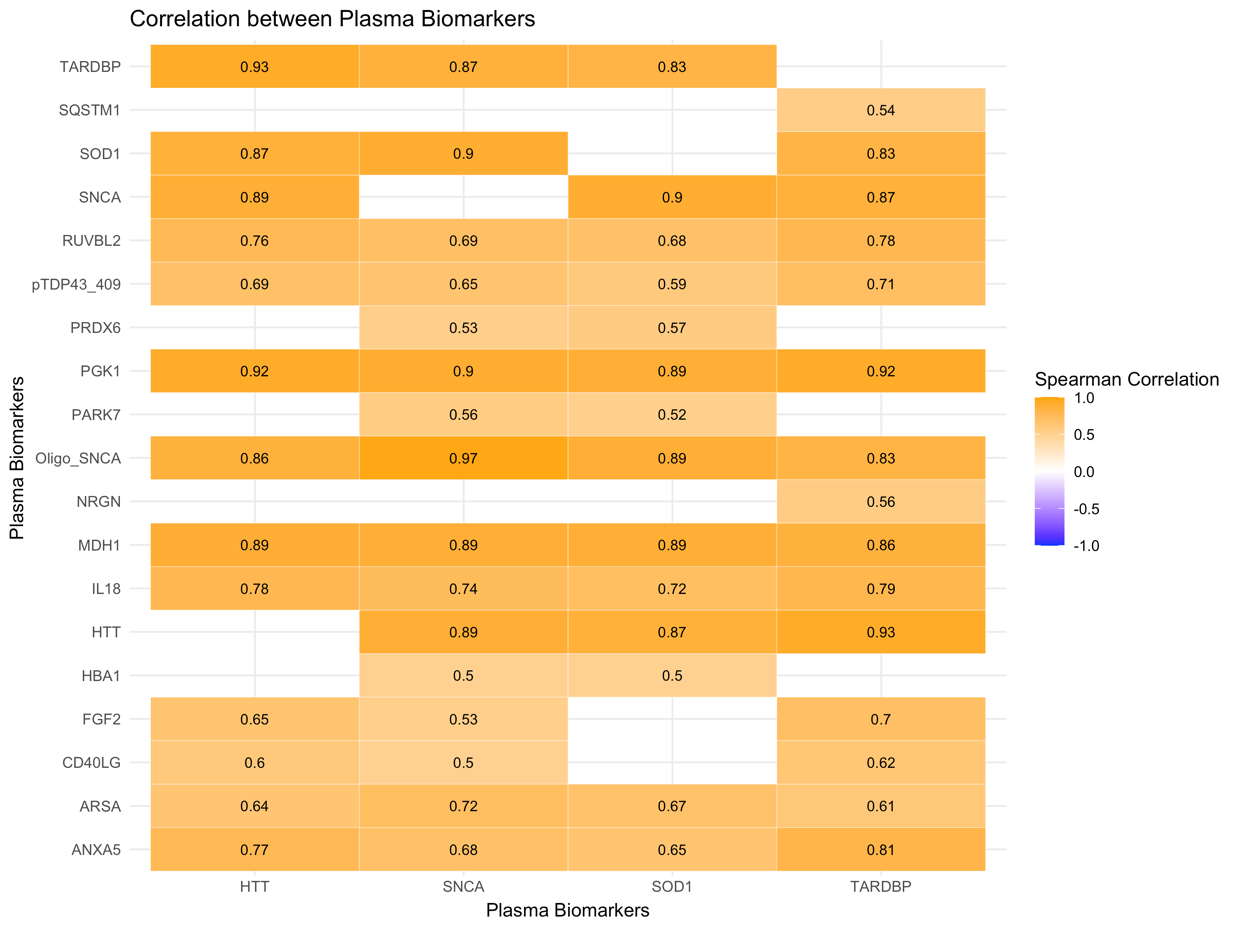

***Supplementary Fig.*** ***4.*** Plasma biomarkers correlation for HTT, SNCA, SOD1 and TARDBP in whole cohort. This heatmap illustrates the Spearman correlation between various plasma biomarkers, highlighting pairwise relationships among markers involved in neurodegeneration. Darker colors indicate higher positive correlations, while lighter shades indicate weaker or no correlation. TARDBP: TAR DNA-binding protein 43, SQSTM1: sequestosome 1, SOD1: superoxide dismutase 1, SNCA: alpha-synuclein, RUVBL2: RuvB-like AAA ATPase 2, pTDP43_409: phosphorylated TAR DNA-binding protein 43 at residue 409, PRDX6: peroxiredoxin 6, PGK1: phosphoglycerate kinase 1, PARK7: parkinsonism associated deglycase (DJ-1), Oligo_SNCA: oligomeric alpha-synuclein, NRGN: neurogranin, MDH1: malate dehydrogenase 1, IL18: interleukin 18, HTT: huntingtin, HBA1: haemoglobin subunit alpha 1, FGF2: fibroblast growth factor 2, CD40LG: CD40 ligand, ARSA: arylsulfatase A, ANXA5: annexin A5

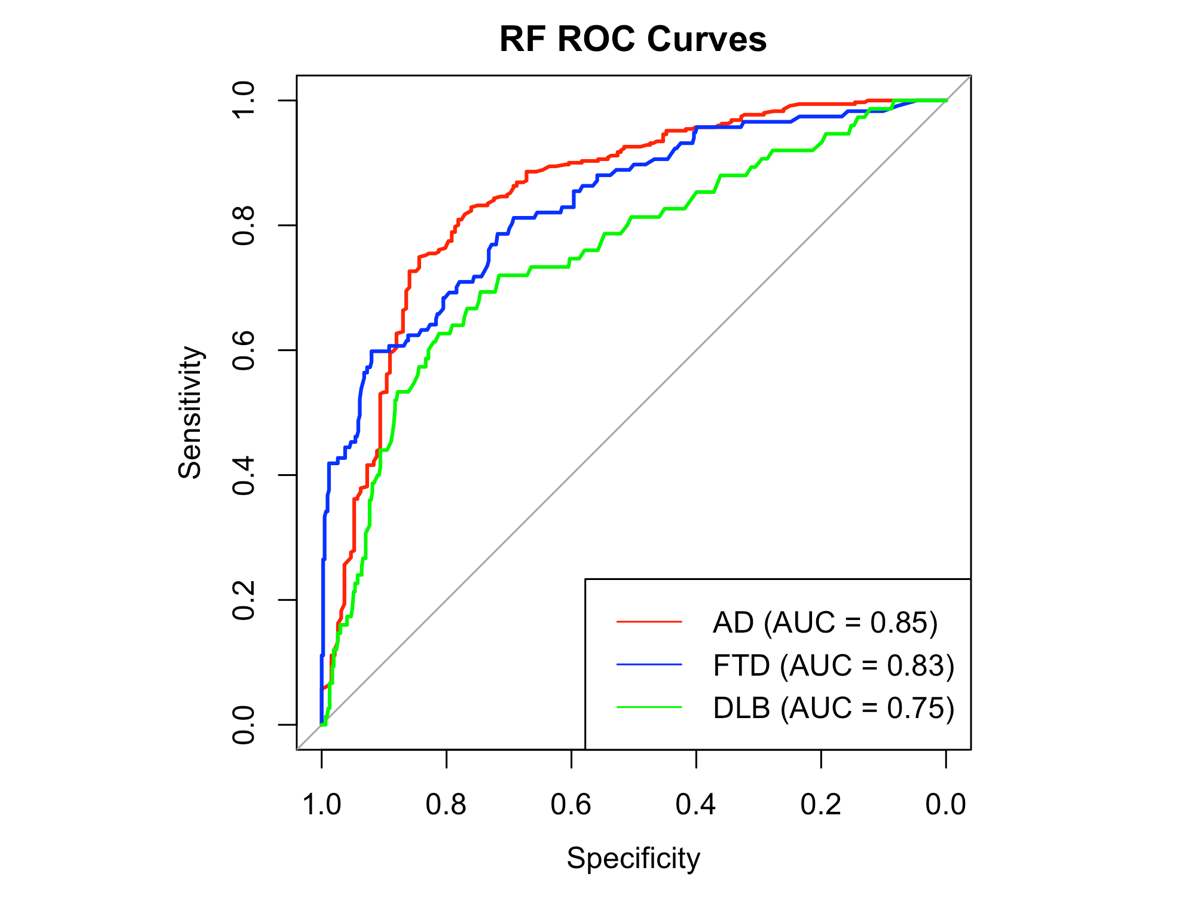

***Supplementary Fig.*** ***5.*** Random forest analysis ROC curve for dementia diagnosis*.* The ROC curves display the diagnostic performance of a model combining all plasma biomarkers, age, and gender for predicting AD, DLB, FTD diagnoses. The AUC for AD, FTD, and DLB are 0.85, 0.83 and 0.75, respectively. AD: Alzheimer's Disease, DLB: Dementia with Lewy Bodies, FTD: Frontotemporal Dementia, AUC: Area Under the Curve

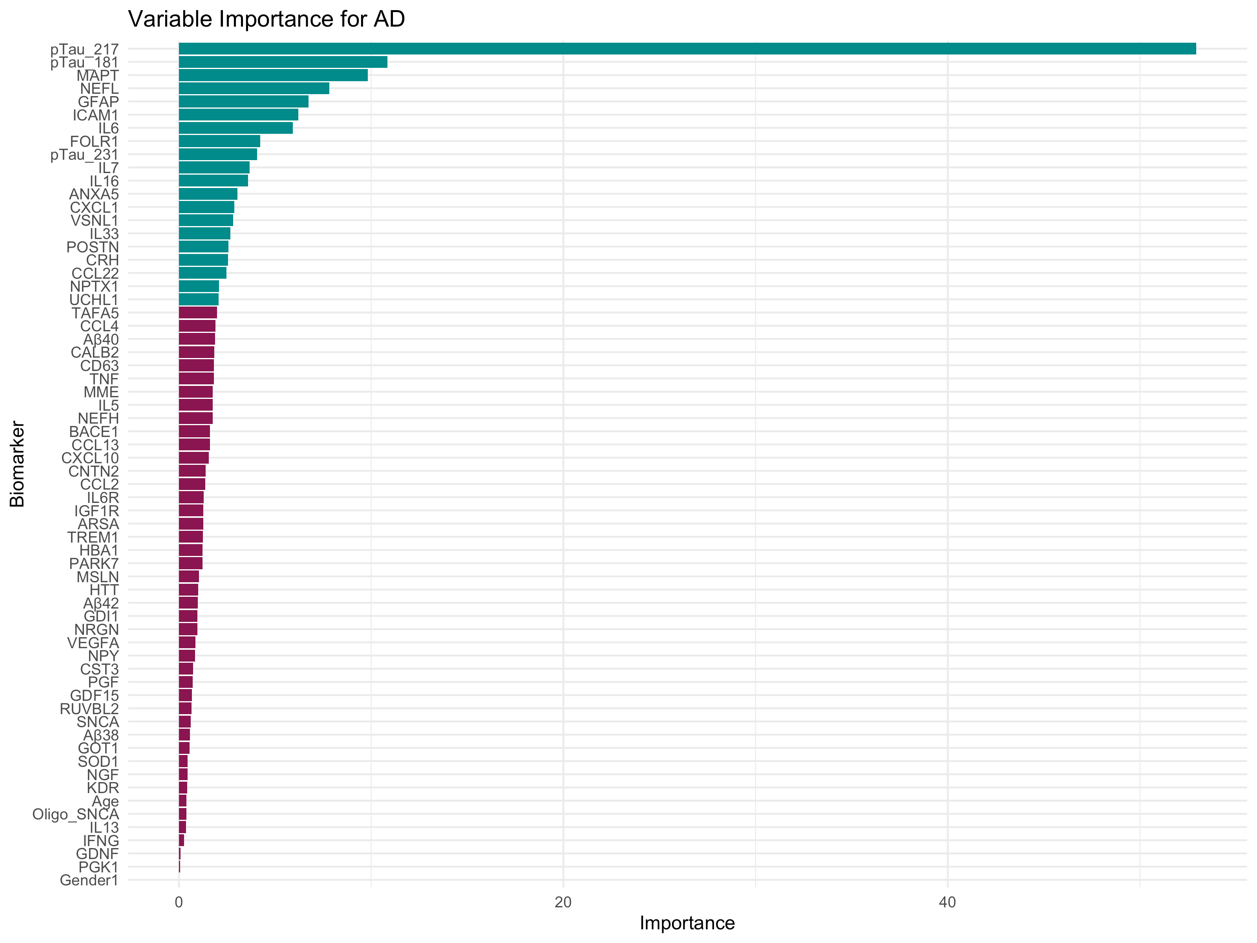

***Supplementary Fig.*** ***6***. Importance of biomarkers for AD diagnosis, based on RF model. This figure ranks biomarkers by their importance in distinguishing Alzheimer’s Disease from other conditions in a random forest model. Biomarkers are ordered by their relative importance score, highlighting the key variables that contribute to the model's predictive accuracy. pTau_217: phosphorylated tau-217, pTau_181: phosphorylated tau-181, MAPT: microtubule-associated protein tau, NEFL: neurofilament light chain, GFAP: glial fibrillary acidic protein, ICAM1: intercellular adhesion molecule 1, IL6: interleukin 6, FOLR1: folate receptor 1, pTau_231: phosphorylated tau-231, IL7: interleukin 7, IL16: interleukin 16, ANXA5: annexin A5, CXCL1: CXC motif chemokine ligand 1, VSNL1: visinin-like protein 1, IL33: interleukin 33, POSTN: periostin, CRH: corticotropin releasing hormone, CCL22: C-C motif chemokine ligand 22, NPTX1: neuronal pentraxin-1, UCHL1: ubiquitin carboxy-terminal hydrolase L1

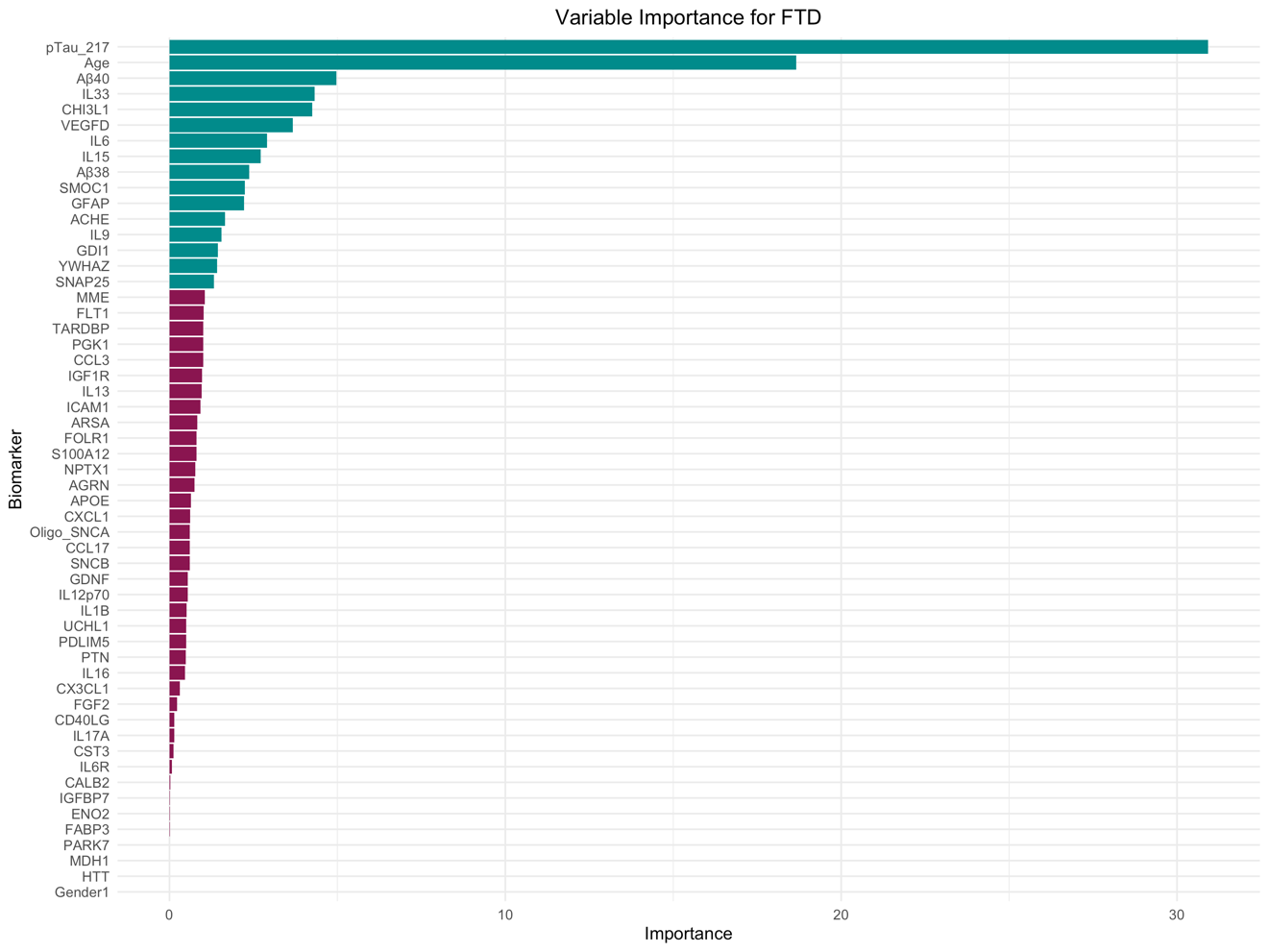

***Supplementary Fig.*** ***7****.* Importance of biomarkers for Frontotemporal Dementia (FTD) diagnosis, based on RF model. This figure ranks biomarkers by their importance in distinguishing FTD from other conditions in a random forest model. Biomarkers are ordered by their relative importance score, highlighting the key variables that contribute to the model's predictive accuracy. pTau_217: phosphorylated tau-21, Aβ40: amyloid-beta 40, IL33: interleukin 33, CHI3L1: chitinase-3-like protein 1, VEGFD: vascular endothelial growth factor D, IL6: interleukin 6, IL15: interleukin 15, Aβ38: amyloid-beta 38, SMOC1: SPARC-related modular calcium-binding protein 1, GFAP: glial fibrillary acidic protein, ACHE: acetylcholinesterase, IL9: interleukin 9, GDI1: GDP dissociation inhibitor 1, YWHAZ: tyrosine 3-monooxygenase activation protein zeta, SNAP25: synaptosomal-associated protein 25.

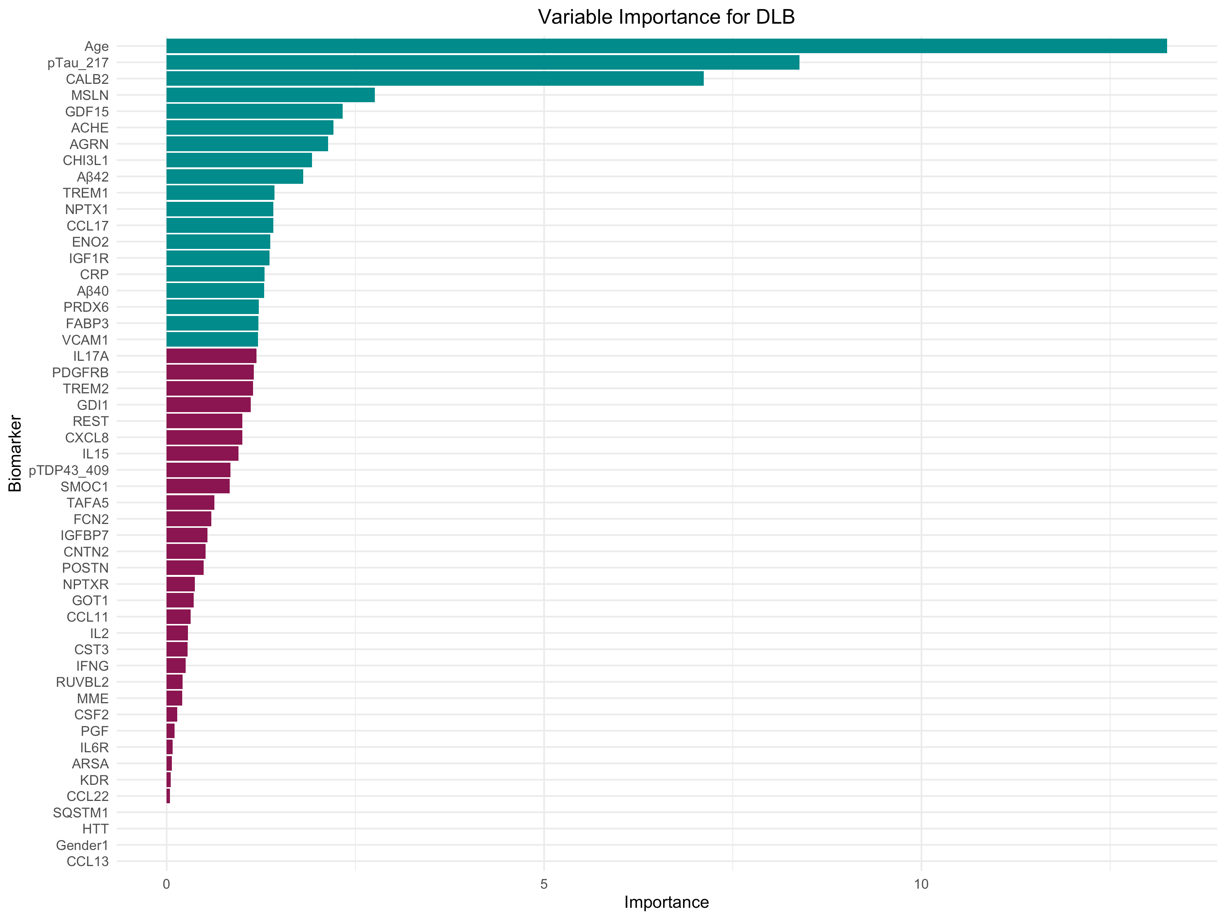

***Supplementary Fig.*** ***8***. Importance of biomarkers for Dementia with Lewy Bodies (DLB) diagnosis, based on RF model. This figure ranks biomarkers by their importance in distinguishing DLB from other conditions in a random forest model. Biomarkers are ordered by their relative importance score, highlighting the key variables that contribute to the model's predictive accuracy. pTau_217: phosphorylated tau-217, CALB2: calbindin 2, MSLN: mesothelin, GDF15: growth differentiation factor 15, ACHE: acetylcholinesterase, AGRN: agrin, CHI3L1: chitinase-3-like protein 1, Aβ42: amyloid-beta 42, TREM1: triggering receptor expressed on myeloid cells 1, NPTX1: neuronal pentraxin 1, CXCL17: C-X-C motif chemokine ligand 17, ENO2: enolase 2, IGF1R: insulin-like growth factor 1 receptor, CRP: C-reactive protein, Aβ40: amyloid-beta 40, PRDX6: peroxiredoxin 6, FABP3: fatty acid-binding protein 3, VCAM1: vascular cell adhesion molecule 1

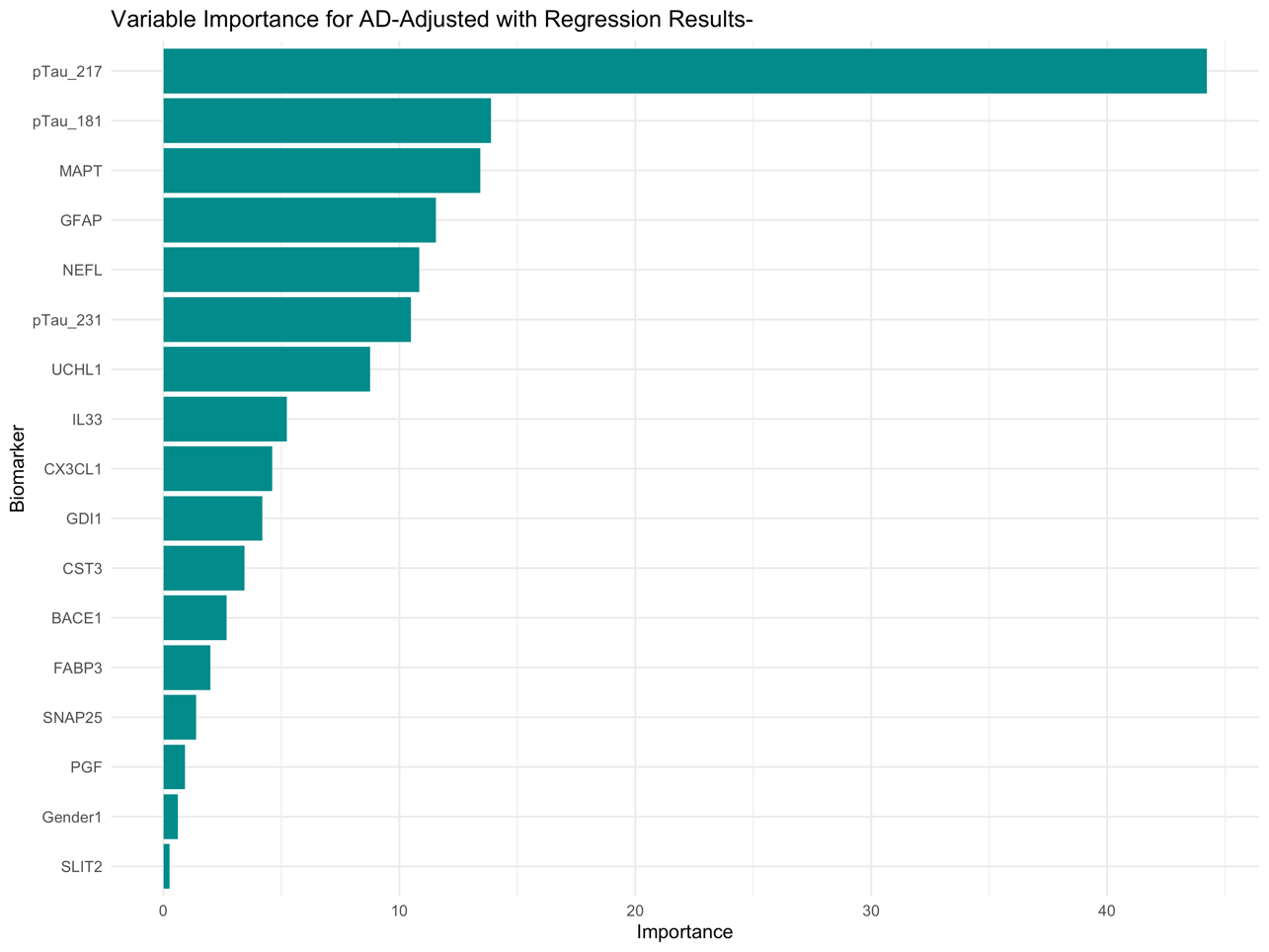

***Supplementary Fig. 9****.* This figure shows the variable importance for distinguishing Alzheimer Dementia (AD), as determined by a Random Forest (RF) model, adjusted with regression results. The bars represent the importance of the top biomarkers in classifying AD patients, based on their Mean Decrease in Gini index. pTau_217: phosphorylated tau-217, pTau_181: phosphorylated tau-181, MAPT: microtubule-associated protein tau, GFAP: glial fibrillary acidic protein, NEFL: neurofilament light chain, pTau_231: phosphorylated tau-231, UCHL1: ubiquitin carboxyl-terminal hydrolase L1, IL33: interleukin-33, CXCL1: CXC motif chemokine ligand 1, GDI1: GDP dissociation inhibitor 1, CST3: cystatin C, BACE1: beta-site amyloid precursor protein cleaving enzyme 1, FABP3: fatty acid-binding protein 3, SNAP25: synaptosomal-associated protein 25, PGF: placental growth factor, SLIT2: slit guidance ligand 2.

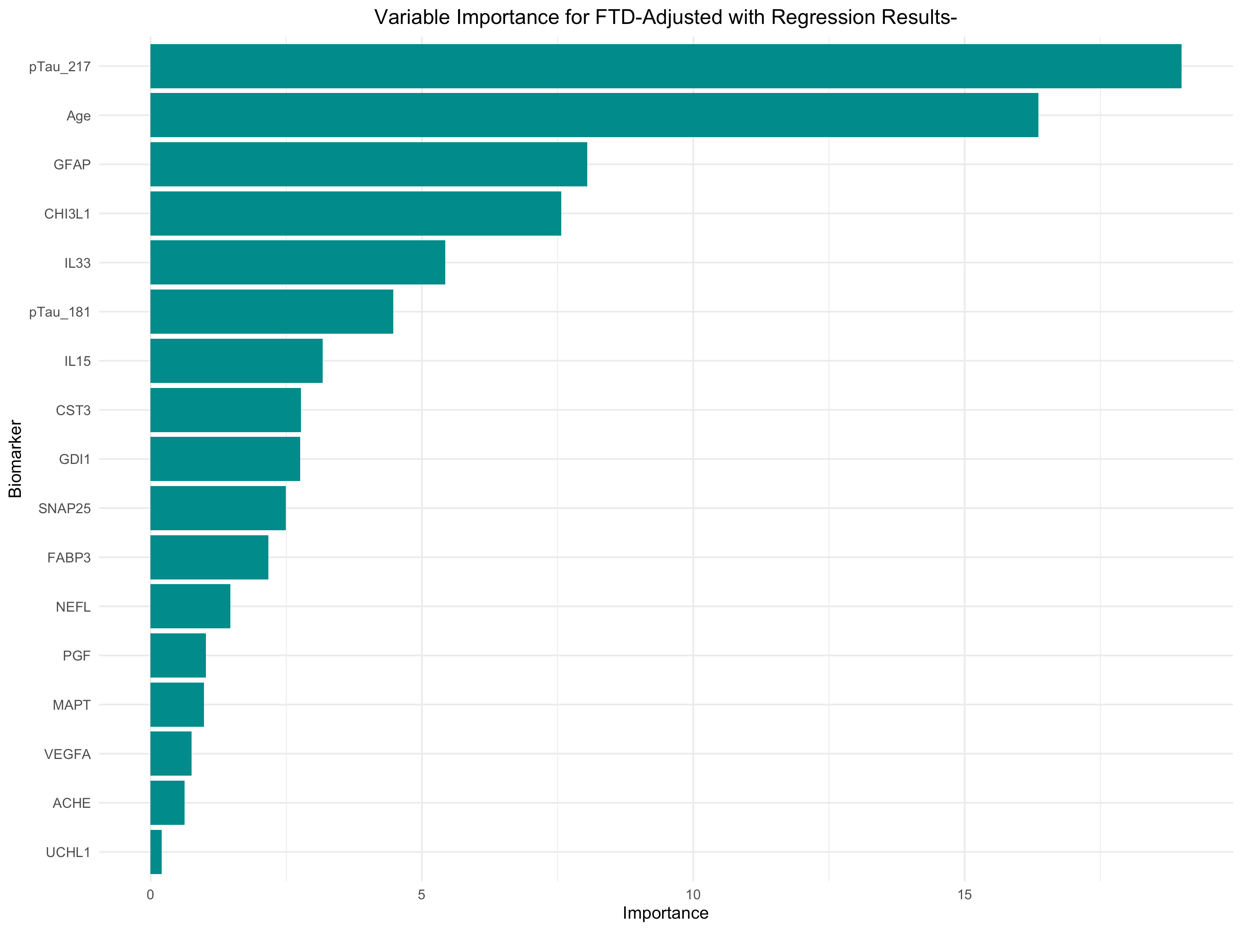

***Supplementary Fig.*** ***10****.* This figure shows the variable importance for distinguishing Frontotemporal Dementia (FTD), as determined by a Random Forest (RF) model, adjusted with regression results. The bars represent the importance of the top biomarkers in classifying FTD patients, based on their Mean Decrease in Gini index. pTau_217: phosphorylated tau-217, GFAP: glial fibrillary acidic protein, CHI3L1: chitinase-3-like protein 1, IL33: interleukin-33, pTau_181:phosphorylated tau-181, IL15: interleukin-15, CST3: cystatin C, GDI1: GDP dissociation inhibitor 1, SNAP25: synaptosomal-associated protein 25, FABP3: fatty acid-binding protein 3, NEFL: neurofilament light chain, PGF: placental growth factor, MAPT: microtubule-associated protein tau, VEGFA: vascular endothelial growth factor A, ACHE: acetylcholinesterase, UCHL1: ubiquitin carboxyl-terminal hydrolase L1.

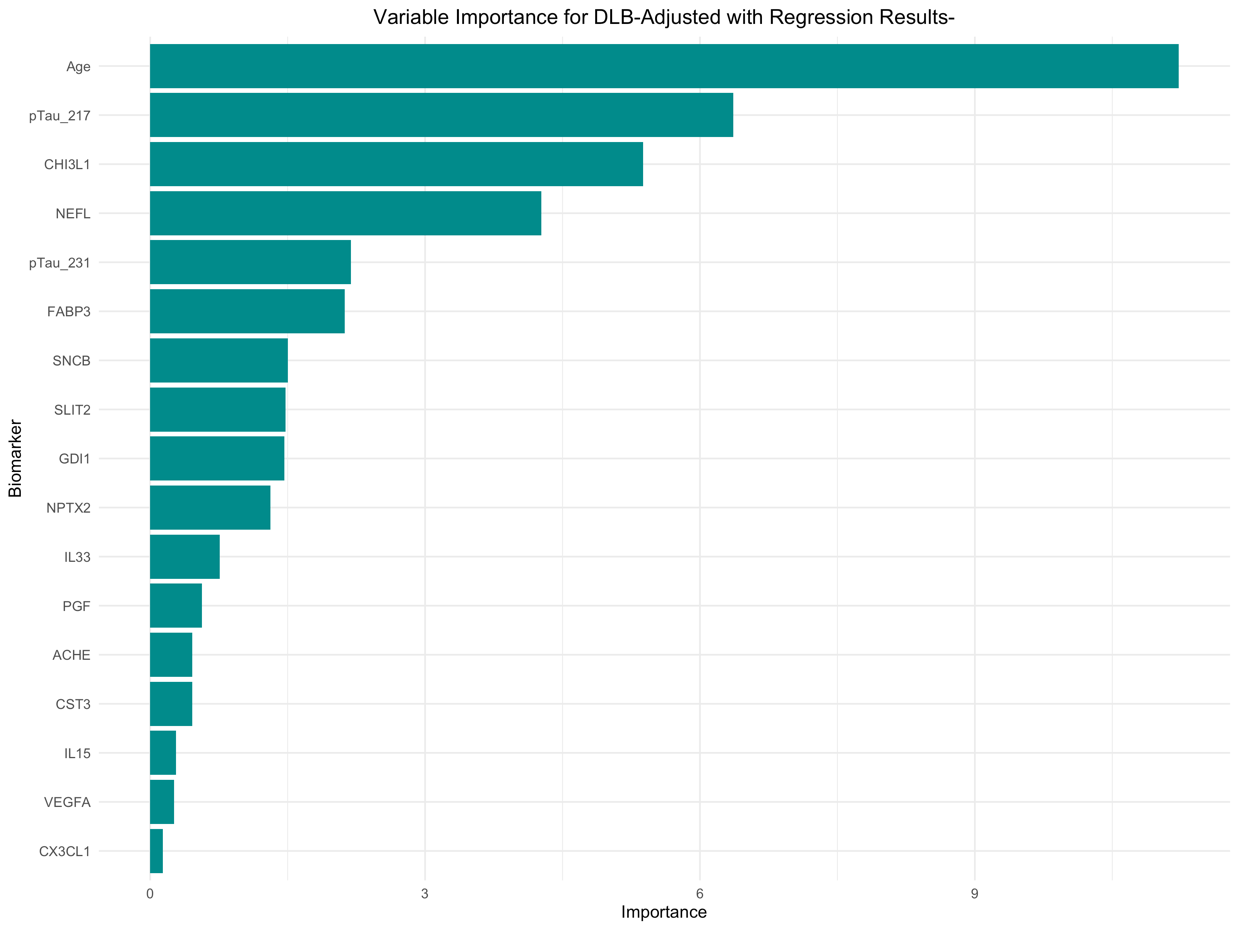

***Supplementary Fig.*** ***11****.* This figure shows the variable importance for distinguishing Dementia with Lewy Bodies (DLB), as determined by a Random Forest (RF) model, adjusted with regression results. The bars represent the importance of the top biomarkers in classifying DLB patients, based on their Mean Decrease in Gini index. Tau_217: phosphorylated tau-217, CHI3L1: chitinase-3-like protein 1, NEFL: neurofilament light chain, pTau_231: phosphorylated tau 231, FABP3: fatty acid-binding protein 3, SNCB: beta-synuclein, SLIT2: slit guidance ligand 2, GDI1: GDP dissociation inhibitor 1, NPTX2: neuronal pentraxin 2, IL33: interleukin-33, PGF: placental growth factor, ACHE: acetylcholinesterase, CST3: cystatin C, IL15: interleukin-15, VEGFA: vascular endothelial growth factor A, CX3CL1: fractalkine.

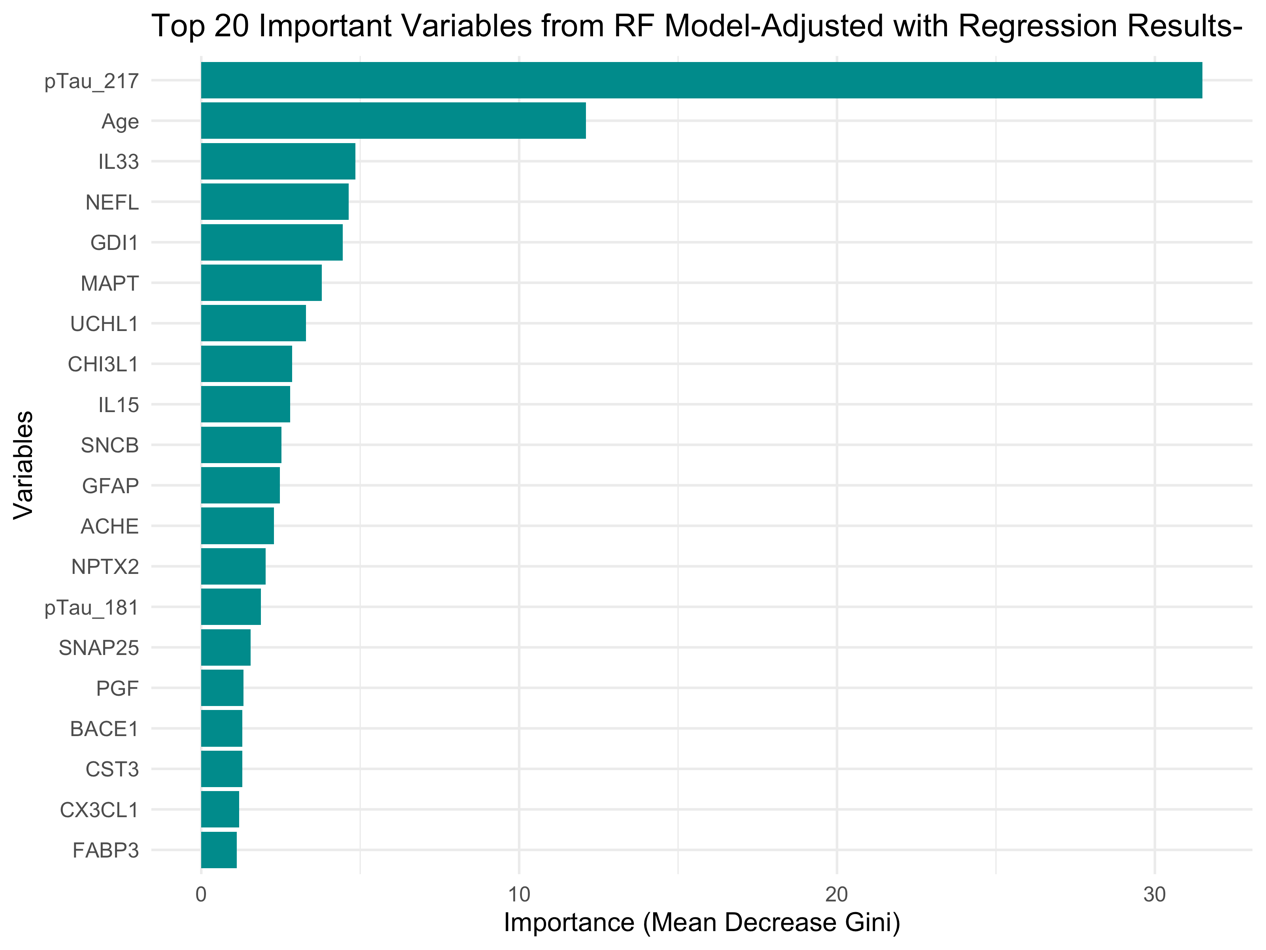

***Supplementary Fig.*** ***12.*** Random forest model’s most important biomarkers for biomarkers that differentiate between dementia types. This figure presents the top 20 important variables identified from a Random Forest (RF) model. The importance of these variables is determined by the Mean Decrease in Gini index, which reflects each variable’s contribution to the accuracy of the classification model. Specifically, this model was adjusted with logistic regression results to improve the robustness of variable selection, focusing on distinguishing between the studied neurodegenerative diseases. pTau_217: phosphorylated tau-217, IL33: interleukin-33, NEFL: neurofilament light chain, GDI1: GDP dissociation inhibitor 1, MAPT: microtubule-associated protein tau, UCHL1: ubiquitin C-terminal hydrolase L1, CHI3L1: chitinase-3-like protein 1, IL15: interleukin-15, SNCB: beta-synuclein, GFAP: glial fibrillary acidic protein, ACHE: acetylcholinesterase, NPTX2: neuronal pentraxin-2, pTau_181: phosphorylated tau-181, SNAP25: synaptosomal-associated protein 25, PGF: placental growth factor, BACE1: beta-site amyloid precursor protein cleaving enzyme 1, CST3: cystatin C, CX3CL1: fractalkine, FABP3: fatty acid-binding protein 3

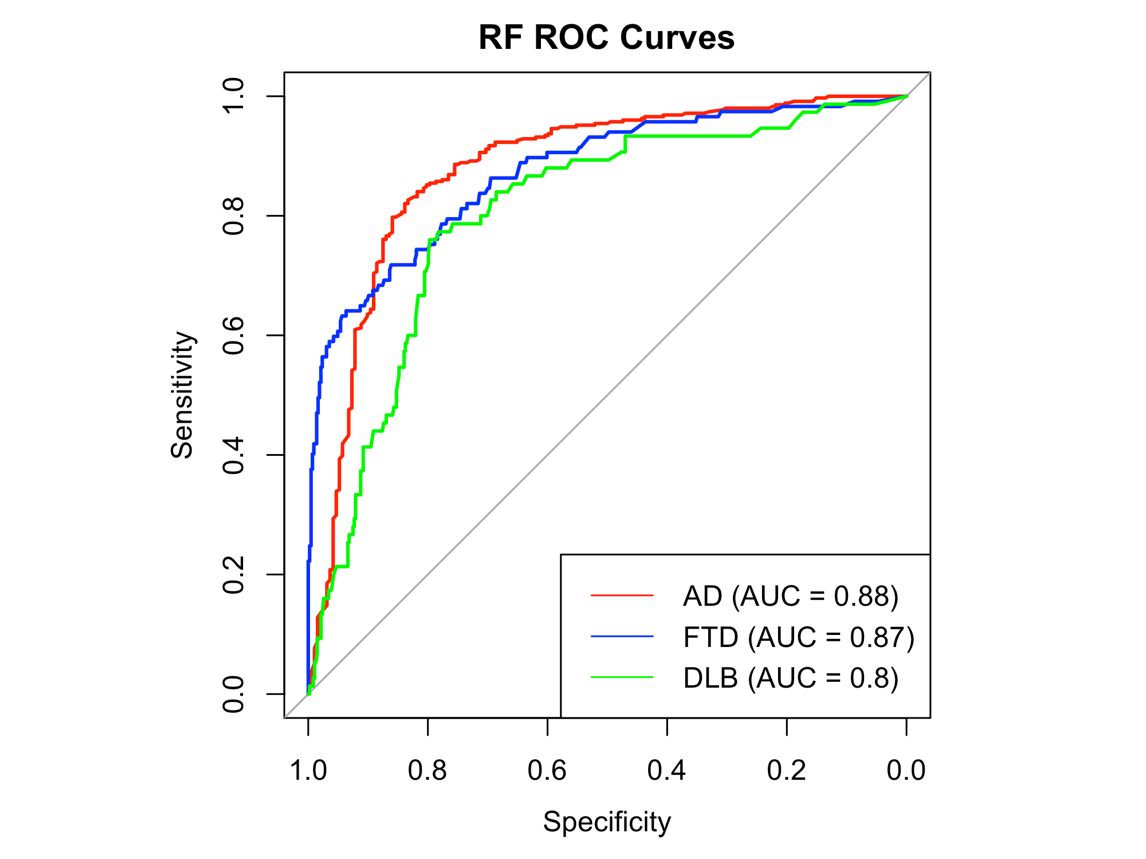

***Supplementary Fig.*** ***13****.* Random forest ROC curve for dementia diagnosis, differentiated biomarkers. The ROC curves display the diagnostic performance of a model combining only differently expressed plasma biomarkers, age, and gender for predicting AD, DLB, FTD diagnoses. The AUC for AD, FTD, and DLB are 0.88, 0.87 and 0.8, respectively. AD: Alzheimer's Disease, DLB: Dementia with Lewy Bodies, FTD: Frontotemporal Dementia, AUC: Area Under the Curve

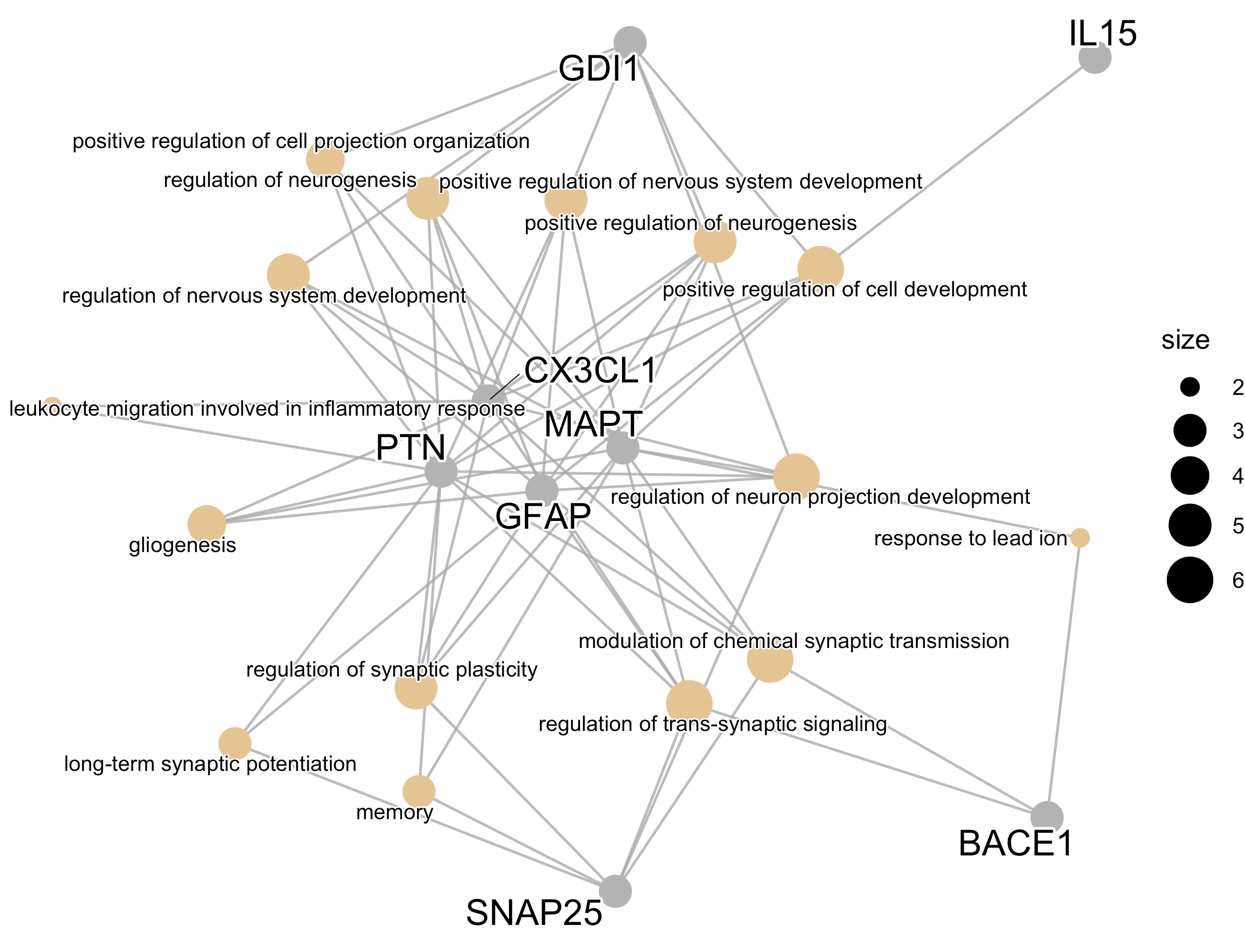

***Supplementary Fig.*** ***14.*** GO enrichment pathway network for differently expressed biomarkers between different dementias. The network plot shows relationships between key biomarkers (gray nodes) such as MAPT (microtubule-associated protein tau), CX3CL1 (C-X3-C motif chemokine ligand 1), GFAP (glial fibrillary acidic protein), SNAP25 (synaptosomal-associated protein 25), BACE1 (beta-site APP cleaving enzyme 1), GDI1 (GDP dissociation inhibitor 1), IL15 (interleukin 15), and PTN (pleiotrophin). Connections represent biological processes (orange nodes) linked to these markers, such as regulation of neurogenesis, synaptic plasticity, gliogenesis, and leukocyte migration involved in the inflammatory response. Node size reflects the relative importance of each biomarker or process, with larger nodes representing more central elements in the network.
